# A Natural History Study of Timothy Syndrome

**DOI:** 10.1101/2024.05.20.24307583

**Authors:** Katherine W. Timothy, Rosemary Bauer, Kerry A. Larkin, Edward P. Walsh, Dominic J. Abrams, Cecilia Gonzalez Corcia, Alexandra Valsamakis, Geoffrey S. Pitt, Ivy E. Dick, Andy Golden

## Abstract

Timothy syndrome (OMIM #601005) is a rare disease caused by variants in the gene *CACNA1C*. Timothy syndrome patients were first identified as having a cardiac presentation of Long QT and syndactyly of the fingers and/or toes, and an identical variant in *CACNA1C*, Gly406Arg. However, since this original identification, more individuals harboring diverse variants in *CACNA1C* have been identified and have presented with various cardiac and extra-cardiac symptoms. Furthermore, it has remained underexplored whether individuals harboring canonical Gly406Arg variants in mutually exclusive exon 8A (Timothy syndrome 1) or exon 8 (Timothy syndrome 2) have additional symptoms. Here, we describe the first Natural History Study for Timothy syndrome, providing a thorough resource describing the current understanding of disease manifestation in Timothy syndrome patients. Parents of Timothy syndrome children were queried regarding a wide-ranging set of symptoms and features via a survey. Importantly, we find that in addition to cardiac concerns, Timothy syndrome patients commonly share extra-cardiac features including neurodevelopmental impairments, hypoglycemia, and respiratory problems. Our work expands the current understanding of the disorder to better inform the care of Timothy syndrome patients.

## INTRODUCTION

In the early 1990s, after a flurry of publications identifying the genes responsible for many long QT (LQT) syndromes^1–5^, several reports were published in which children were identified as having a prolonged QT interval and syndactyly^6–9^. At the time, Katherine Wilson Timothy (KWT) was the Clinical Coordinator for Dr. Mark T. Keating’s genetic research laboratory at the University of Utah which was searching the genome for variants that caused a prolonged QT interval and other cardiac abnormalities associated with sudden unexplained death (SUD) in the young. Recognizing these studies, cardiologists referred children to the laboratory with a specific LQT + syndactyly phenotype. Data from these children were collected, as this was believed to be a new syndrome. With 17 such children identified, most from the United States, Italy, New Zealand and the United Kingdom, the identity of the shared culprit gene was finally discovered after more than ten years of searching: *CACNA1C,* encoding the α_1C_ subunit of the voltage-gated L-type calcium channel Ca_v_1.2, which represents the dominant voltage-gated calcium channel within the heart. Thirteen of the 17 individuals had the identical missense variant, Gly406Arg, but DNA was not analyzed for the remaining four of these phenotypically identical children. Most children acquired this variant *de novo*^10^. The cardiac phenotypes presented by this autosomal dominant syndrome were thought to be the result of failed inactivation of Ca_v_1.2. Given that it was the eighth LQT gene to be identified molecularly, the LQT in these individuals was given the name LQT8. The syndrome was named Timothy syndrome (TS) (OMIM #601005) in recognition of KWT’s work. While TS was first identified due to shared LQT and syndactyly phenotypes among patients, it has become clear that these individuals experience variable symptoms across multiple organ systems. Thus, a systematic investigation into what phenotypes occur in TS is critical for proper identification and treatment of the disease.

The *CACNA1C* gene has at least 47 major exons and numerous splice variant products^11, 12^. An important difference between known splice forms is their inclusion of either exon 8 or 8A. Exons 8 and 8A are identical in size, and individual messenger RNAs derived from the *CACNA1C* gene either incorporate sequences from either exon 8 or 8A, but never both. Thus, we refer to them as being mutually exclusive.

The 13 original TS children had the identical variant in *CACNA1C* exon 8A, resulting in a missense change Gly406Arg^10^. This was originally defined as the cause of TS, later designated as TS type 1 (TS1), and was the first reported disease variant in *CACNA1C*. A single case of TS (but without syndactyly) occurring in the alternative exon 8 was subsequently identified, which was then designated as TS type 2 (TS2)^13^. This second variant resulted in the homologous missense change Gly406Arg. In the literature, the TS1 and TS2 types are often referred to as synonymous syndromes, mainly due to the identified similarities in their cardiac phenotypes. However, the development of many of the other systems (e.g., neuronal, bone, skeletal and smooth muscle development) are recognized to be differently affected. Exons 8 and 8A are used at distinct times and are dynamically regulated during mouse and human cerebral cortical development^14^. Similar regulation may occur in other organs or tissues. TS variants often arise de novo. The timing of occurrence can cause mosaicism leading to differential expression of mutant and wild type Cav1.2 channels within specific tissues, complicating the comparison between TS1 and TS2 and among TS1 or TS2 affected individuals.^15, 16^.

An additional variant, Gly402Ser in exon 8, was identified in a separate individual concurrently with the TS2 change in 2005^13^. It was reported in one individual who had LQT and neurological symptoms, but no syndactyly. Despite the different disease presentation from those with Gly406Arg, this case was classified as TS2 due to the phenotypic understanding at the time and the simultaneous discovery. In this study, Gly402Ser was analyzed separately from Gly406Arg TS1 and TS2 to enable comparison between the variants and the identification of unique phenotypes associated with a particular variant.

Numerous additional variants in the *CACNA1C* gene have been identified. Individuals with *CACNA1C* variants who have LQT but no immediately identified extra-cardiac symptoms typically received a diagnosis of LQT8. For the purpose of this study, we will separate these individuals and refer to them as diagnosed with non-syndromic LQT8 (nsLQT8). Individuals who have some symptom overlap with TS1 and TS2 have received a variety of diagnoses, including atypical TS, and *CACNA1C*-related disorder. While many of these individuals have been described in detail as individual case reports, we aim to analyze them together to highlight the many similarities in their disease presentation.

## METHODS

### Inclusion

KWT has been the face of TS since it was first recognized as a syndrome and for the past three decades has served as the main point of contact for physicians and families when a child is first diagnosed. Continuous contact with many TS families has been maintained through in-person visits, attendance at annual Sudden Arrythmia Death Syndromes (SADS) and Timothy Syndrome Foundation Conference meetings, social media, email, phone calls, and letter writing. For this reason, we were able to invite 105 individuals to participate in this Natural History Study, having previously collected and retained their contact information (Figure 1).

**Figure 1.**
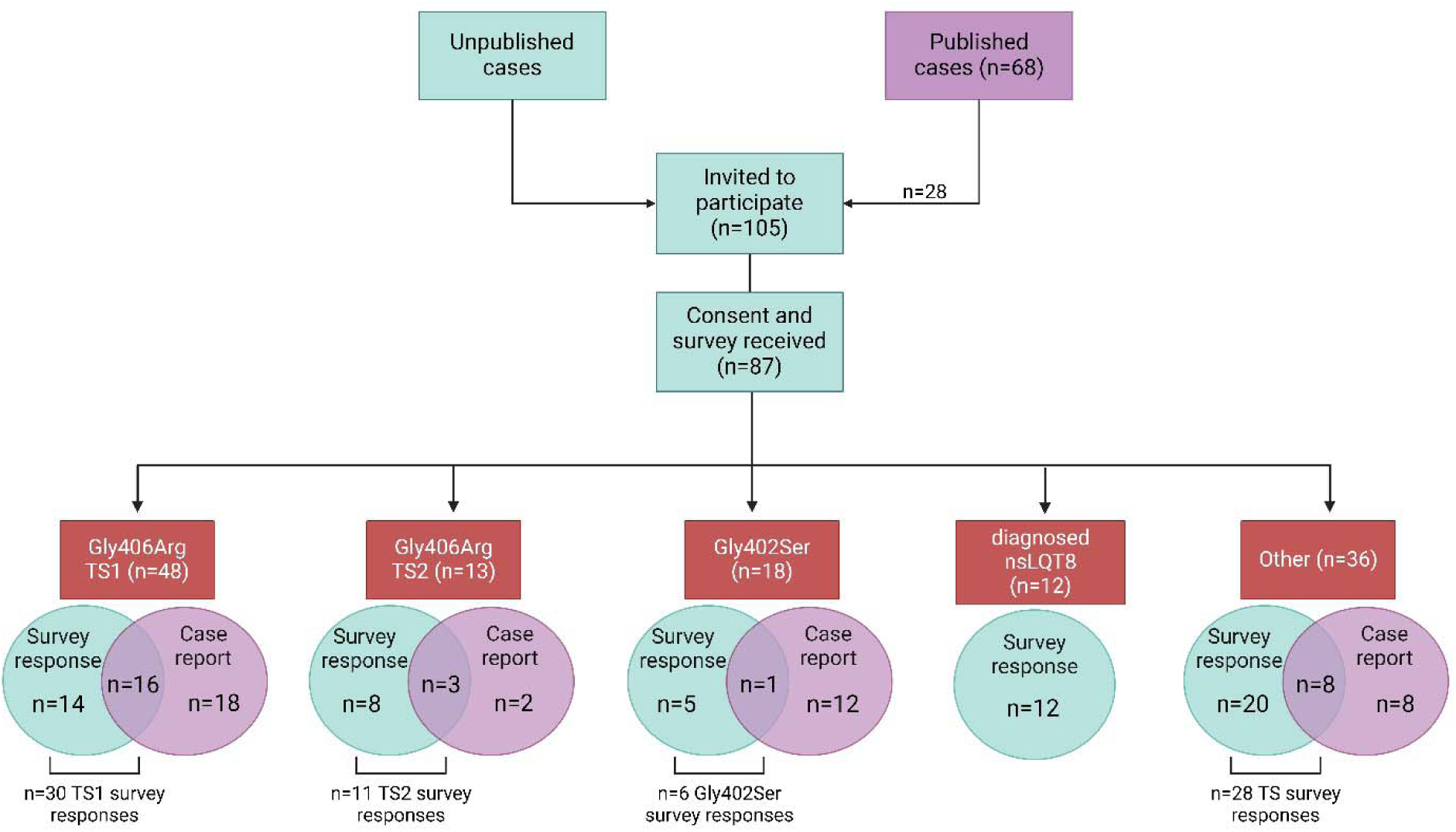
Schematic of the study participants. Blue indicates survey respondents, which comprise all data presented within the tables. Purple indicates cases published in the literature, with overlap of survey respondents as shown. Image created with BioRender.com.

Study participation required the presence of a *CACNA1C* variant and symptoms overlapping the original features of TS. In most individuals, a variant was identified using a cardiac gene panel (typically 40-50 genes); contributions of potential variants in other genes cannot be assessed. Whole exome analysis was performed in a few cases.

Case counts and relative proportions of each variant were obtained from published (n=52) and unpublished individuals (Figure 1). To avoid duplication of case counts, survey respondents were asked to report whether their case was published and to include the citation. If it was unclear whether a case was a duplicate, direct contact was made with contributing authors. Symptom occurrence reported in the tables herein included only data obtained from survey responses because of the relative incompleteness of information provided in case reports, as these publications focus mostly on cardiac presentations of neonates and generally do not include any information on extra-cardiac symptoms or the constellation of symptoms that may present later in childhood.

In order to facilitate comparison across reportedly different phenotypes, the study participants were organized into 5 separate categories (Figure 1). *A*: 30 survey participants reported the Gly406Arg variant in exon 8A, comprising the TS1 group. *B*: 11 survey participants reported the Gly406Arg variant in exon 8. This group is identified as TS2 Gly406Arg. *C*: 6 survey participants harbored the Gly402Ser variant in exon 8. While this group was historically included within TS2, we choose to analyze these patients separately to compare and contrast the effects of distinct amino acid changes vs. exon locus. *D:1*2 survey respondents reported a diagnosis of LQT8 only, with a variety of distinct *CACNA1C* variants across this group. In order to evaluate whether these patients truly represent a cardiac-selective phenotype, we group these participants as diagnosed nsLQT8. E: The final category represents all participants who reported a *CACNA1C* variant but do not fall within one of the other four categories (28 survey respondents), Among these, no more than two participants reported the same amino acid change.

### Survey

To characterize the natural history of TS, a 150-question survey was administered via Google Forms to collect information on symptoms associated with organ systems known to be affected, treatments, and outcome. The survey also included questions about pregnancy and prenatal concerns in the mothers. The majority of respondents were the parents/guardians who completed the survey on behalf of their TS children; several adults (largely individuals diagnosed with nsLQT8) answered for themselves. No medical records were requested. Responses were collected after informed consent and the study was approved by the Institutional Review Board of Genetic Alliance (Washington, DC, USA).

Not all questions were answered by each participant. We did not assume that a non-answer was the equivalent of “No,” and thus the sample size for each question reflects the total number of respondents for that question.

## RESULTS

### Study population

#### Overall survey yield

Of the 105 individuals invited to participate in the survey, 87 submitted a consent form and completed the survey. Of those 87, 28 had also been published previously as case reports that were focused primarily on cardiac presentations (Figure 1). These individuals were able to provide additional information on extra-cardiac phenotypes via the survey. Affected individuals represented in the survey data range from 1-71 years of age, with an average age of 13.8 among 71 living individuals. Many of the surveyed individuals carried *CACNA1C* gene variants different from the canonical TS1 and TS2 variants, but nonetheless shared some TS phenotypes. Several patients also contain a frameshift or deletion in *CACNA1C* rather than a missense variant (Table 2: Genotypes).

**Table 1:**
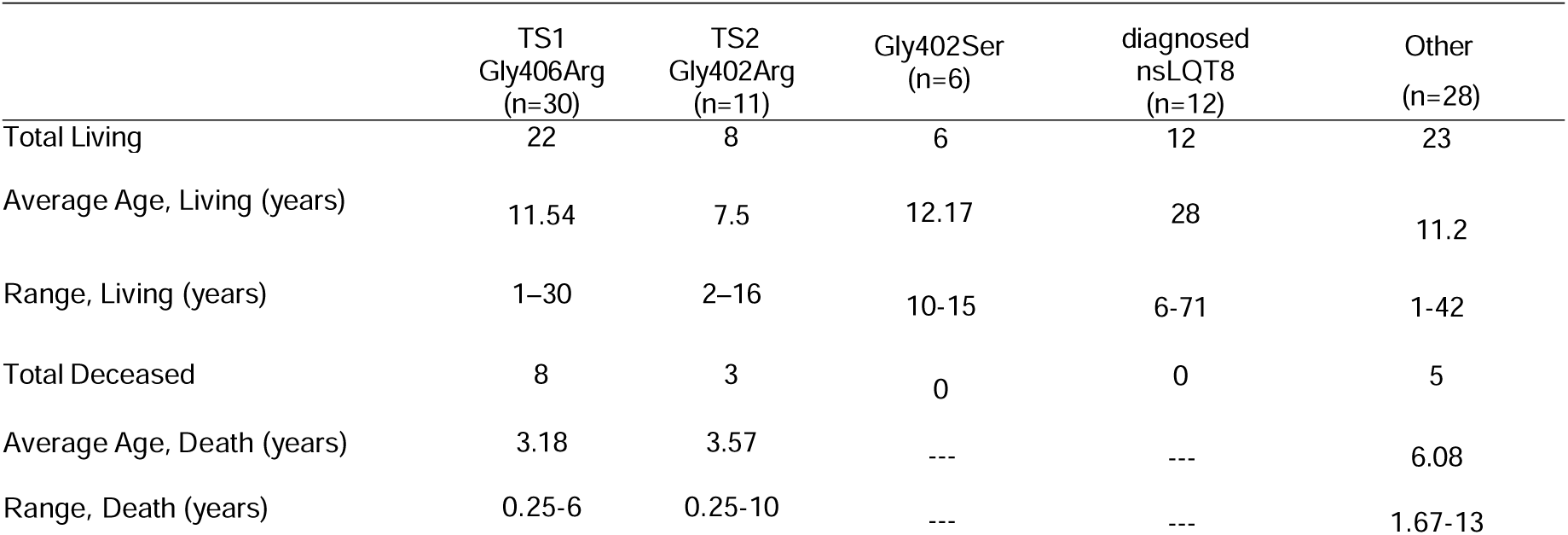
Average Age of Death and Surviving Cohort.

**Table 2:**
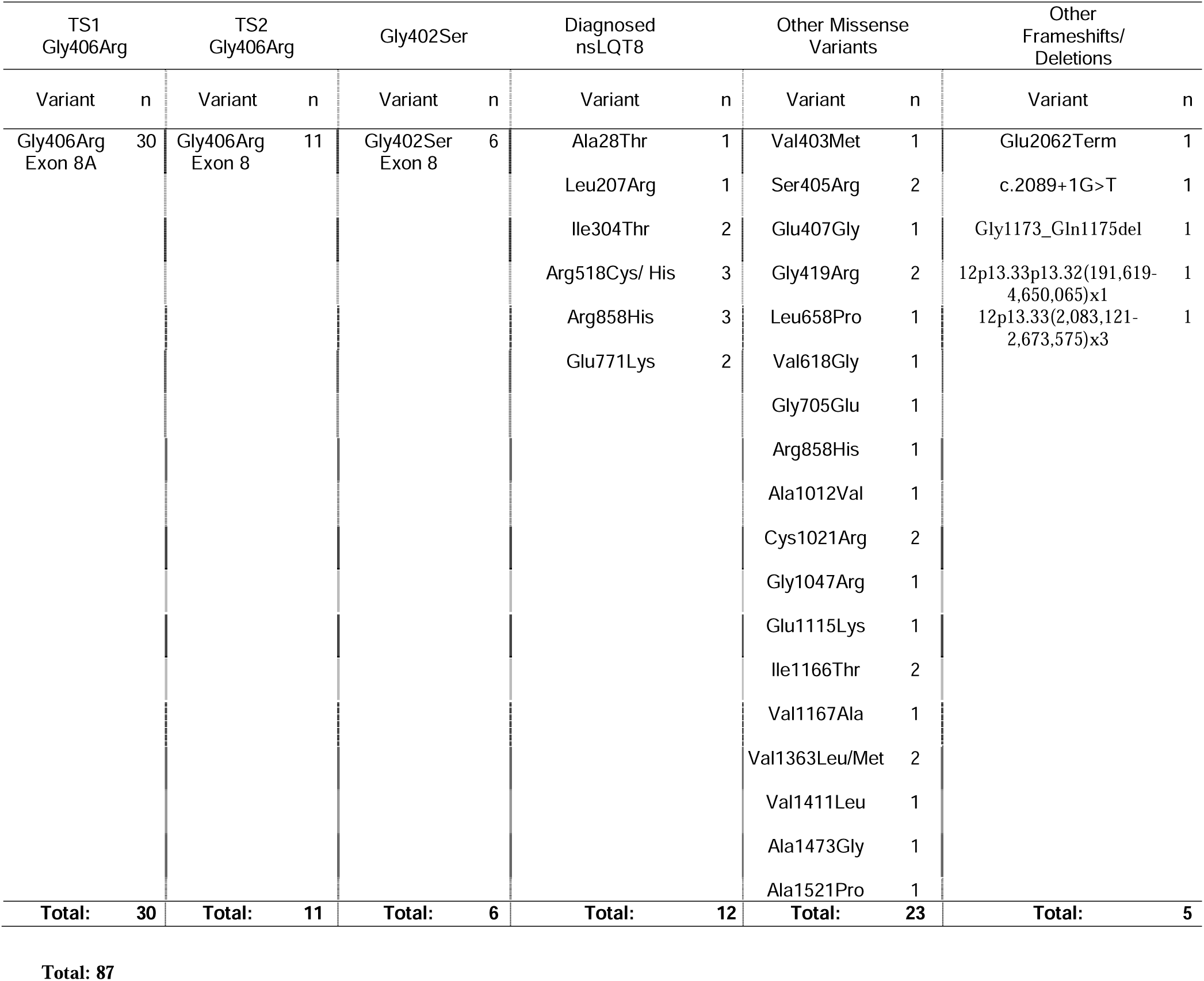
Genotypes:

| TS1<br>Gly406Arg |  | TS2<br>Gly406Arg |  | Gly402Ser |  | Diagnosed<br>nsLQT8 |  | Other Missense<br>Variants |  | Other<br>Frameshifts/<br>Deletions |  |
| --- | --- | --- | --- | --- | --- | --- | --- | --- | --- | --- | --- |
| Variant | n | Variant | n | Variant | n | Variant | n | Variant | n | Variant | n |
| Gly406Arg<br>Exon 8A | 30 | Gly406Arg<br>Exon 8 | 11 | Gly402Ser<br>Exon 8 | 6 | Ala28Thr | 1 | Val403Met | 1 | Glu2062Term | 1 |
|  |  |  |  |  |  | Leu207Arg | 1 | Ser405Arg | 2 | c.2089+1G>T | 1 |
|  |  |  |  |  |  | Ile304Thr | 2 | Glu407Gly | 1 | Gly1173_Gln1175del | 1 |
|  |  |  |  |  |  | Arg518Cys/ His | 3 | Gly419Arg | 2 | 12p13.33p13.32(191,619-4,650,065)x1 | 1 |
|  |  |  |  |  |  | Arg858His | 3 | Leu658Pro | 1 | 12p13.33(2,083,121-2,673,575)x3 | 1 |
|  |  |  |  |  |  | Glu771Lys | 2 | Val618Gly | 1 |  |  |
|  |  |  |  |  |  |  |  | Gly705Glu | 1 |  |  |
|  |  |  |  |  |  |  |  | Arg858His | 1 |  |  |
|  |  |  |  |  |  |  |  | Ala1012Val | 1 |  |  |
|  |  |  |  |  |  |  |  | Cys1021Arg | 2 |  |  |
|  |  |  |  |  |  |  |  | Gly1047Arg | 1 |  |  |
|  |  |  |  |  |  |  |  | Glu1115Lys | 1 |  |  |
|  |  |  |  |  |  |  |  | Ile1166Thr | 2 |  |  |
|  |  |  |  |  |  |  |  | Val1167Ala | 1 |  |  |
|  |  |  |  |  |  |  |  | Val1363Leu/Met | 2 |  |  |
|  |  |  |  |  |  |  |  | Val1411Leu | 1 |  |  |
|  |  |  |  |  |  |  |  | Ala1473Gly | 1 |  |  |
|  |  |  |  |  |  |  |  | Ala1521Pro | 1 |  |  |
| **Total:** | **30** | **Total:** | **11** | **Total:** | **6** | **Total:** | **12** | **Total:** | **23** | **Total:** | **5** |

#### Participants with TS1

Survey responses were obtained for 30 individuals with TS1 (14 female and 16 male), 16 of whom had been identified previously in case reports and 14 who were not previously reported (Figure 1). We also identified 18 case reports of genetically-confirmed individuals, and 4 non-confirmed cases included in the Splawski et al., 2004 publication^10^, for which no parental surveys were acquired. Hence, 48 genetically-confirmed Gly406Arg (exon 8A) individuals were identified, 34 of whom included in this study were previously reported (some individuals were included in more than one published report)^10, 15–29^.

#### Participants with TS2

Survey responses were obtained from 11 individuals with TS2 who harbor the Gly406Arg variant within exon 8 (5 female and 6 male), including 3 individuals previously identified in case reports^13, 25, 30, 31^. Two additional individuals were reported solely in the literature but did not complete a survey^13, 31^, resulting in 13 individuals known to have TS2 (Gly406Arg, Exon 8) (Figure 1).

Another variant, Gly402Ser (exon 8), was originally classified as TS2 in a single child^13^. We collected 6 surveys from individuals with this variant. All 6 cases in our cohort are living. However, as more individuals have been identified with this specific variant, it has become clear that this variant produces a distinct phenotype from TS2 Gly406Arg. We therefore describe this variant separately.

#### Participants with additional variants in CACNA1C

We collected surveys for 40 individuals with a range of molecular changes in *CACNA1C* (Figure 2). One group was a cohort of individuals diagnosed with only LQT8. There is extensive literature on *CACNA1C* variants that selectively cause LQT8 [for review, see ^32^] that suggests individuals with these variants may not have extra-cardiac phenotypes common to individuals diagnosed with TS. In our survey, nine nsLQT8 families encompassing 12 individuals responded. We analyzed these individuals as a separate group, designated ‘diagnosed nsLQT8’, in order to evaluate the hypothesis that these patients do not harbor any extra-cardiac symptoms. Our analysis includes only variants noted in survey responses, although additional variants have been reported in the literature. (Table 2: Genotypes)

**Figure 2.**
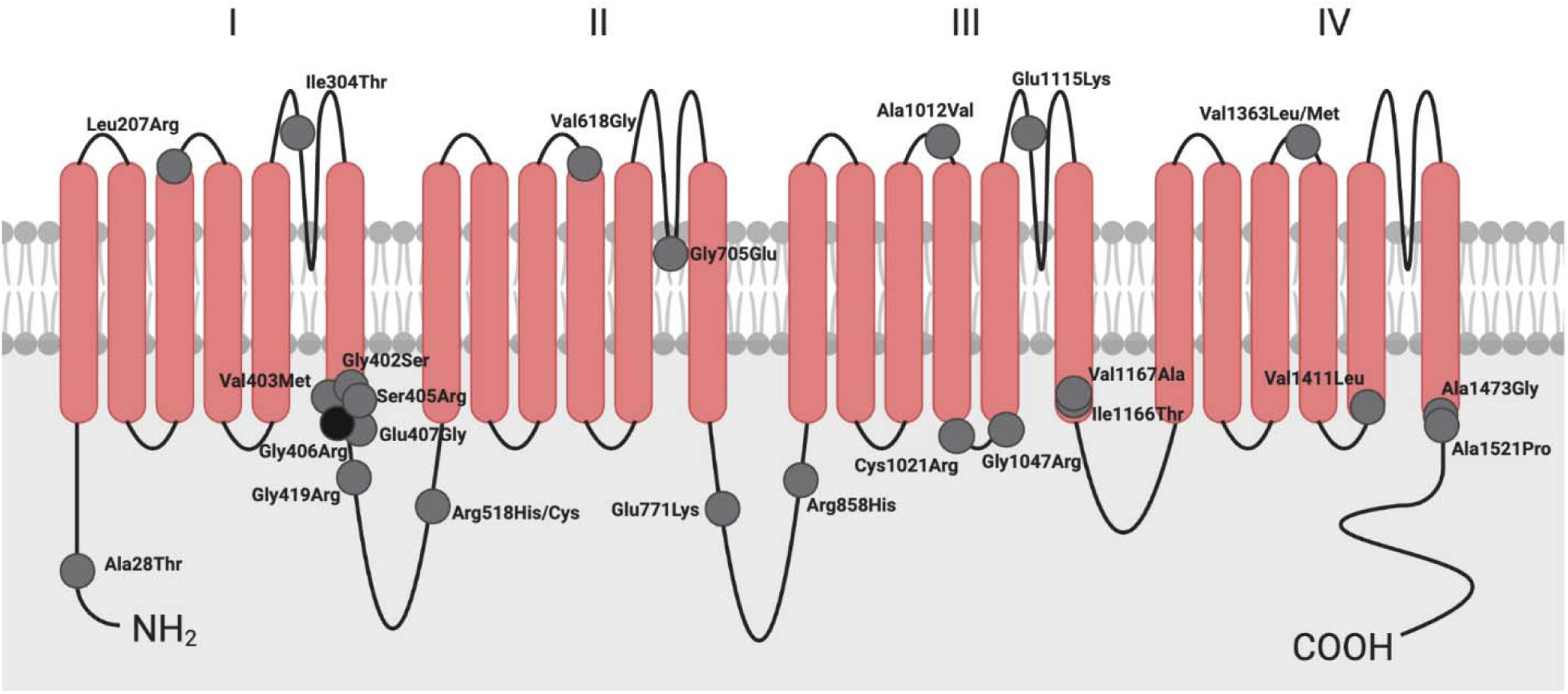
Schematic of the Ca_V_1.2 pore forming subunit showing the locus of mutations described within this study. Image created with BioRender.com.

The remaining 28 respondents with additional variants in *CACNA1C* carry a variety of diagnoses, such as atypical TS or *CACNA1C*-related disorder. In the present study, we analyze these respondents as a single combined group.

#### Average cohort ages & average ages of deaths

The original 2004 TS phenotypic characterization reported the known average age of death of 17 TS1 children at 2.5 years^10^. Survival ages were not reported in that study because of the small number and young ages of the children. Of the 18 literature-reported TS1 children, nine of 18 (50%) died by the time of publication of their respective reports. The calculated average age of these combined literature-reported deaths was 1.13 years, lower than the original average age of death, possibly due to earlier diagnosis. For the other nine literature-reported individuals, survival status was not reported.

Of the thirty TS1 children in our cohort, 8/30 (27%) have died, with an average age at death of 3.2 years (age range of death 3 months-6 years). Seven of these eight were male children. The average age of living participants is 11.54 years (age range 1-30 years). The average age of living males is 14 years (range 5-22 years) and of females, 13 years (range 1-30 years) (Table 1: Average Age of Death and Surviving Cohort).

The original description of TS2 reported on one TS2 Gly406Arg child who died after publication before the age of 10^13^. A second report described a TS2 Gly406Arg child who died in infancy^31^. Of the 11 TS2 Gly406Arg individuals for whom surveys were obtained, three have died between the ages of 3 months and 10 years. The average age of the surviving 8 TS2 Gly406Arg children in our cohort is 7.5 years (age range 2-16 years) (Table 1: Average Age of Death and Surviving Cohort).

Individuals with the Gly402Ser variants in our study averaged 12.17 years of age, ranging from 10-15 years and did not include any individuals who were deceased. In all cases for which exon locus was reported, the variant occurred within exon 8, corresponding to the original description of TS2. All individuals with a nsLQT8 diagnosis were living, averaging 28 years of age (range 6-71 years). Finally, for other *CACNA1C* variants, the 23 living individuals averaged 11.2 years of age (range 1-42 years) 5 individuals died at ages averaging 6.08 years at death (range 1.67-13 years).

### Cardiac Presentations from Gestation through Early Childhood

#### Prenatal presentations

In our TS1 cohort, 13/30 (43%) fetuses had cardiac presentations *in utero,* all presenting with fetal bradycardia. In 8 of these patients (8/13, 62%), the bradycardia was identified as secondary to 2:1 atrioventricular block (AVB). The earliest recognition of these arrhythmias was at 19 weeks gestation (Table 3: Early Development – Cardiac Concerns).

**Table 3:**
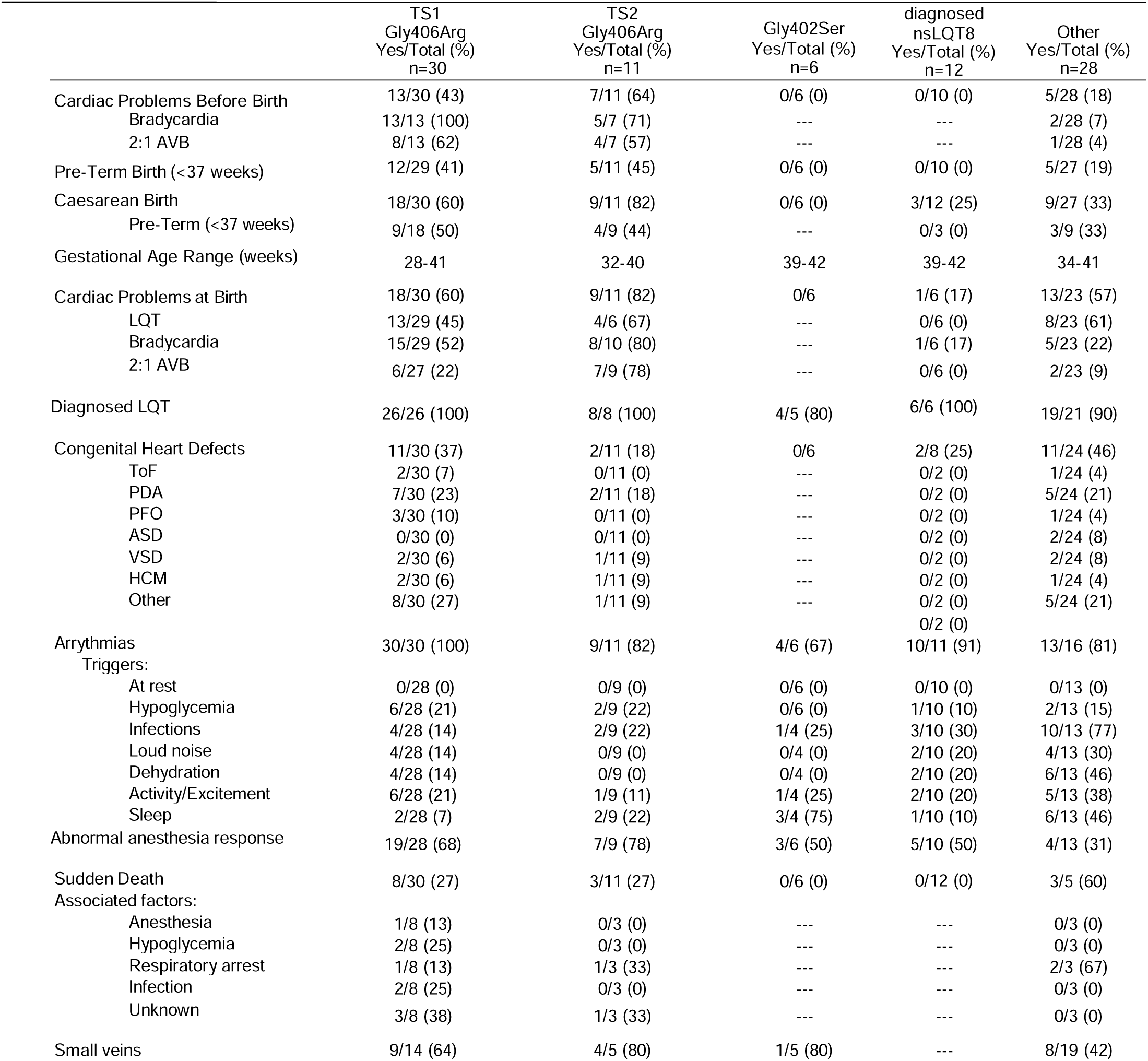
Early Development – Cardiac Concerns.

Caesarean births were frequent among pregnancies with TS1 fetuses, occurring in 18/30 (60%), with a range of fetal gestational age at birth of 28-41 weeks. Of these caesarean births, half were delivered pre-term (before gestation week 37) (9/18, 50%). Three additional pre-term TS1 births occurred via vaginal delivery.

Electrocardiographic abnormalities were present at birth in 18/30 (60%) of TS1 newborns (13 LQT, 15 bradycardia, and 6 with 2:1 AVB). Arrhythmias were detected later in infancy or childhood in additional children; 9 had arrhythmias during surgery, and 2 (18%) suffered unprotected cardiac arrest before age 12, prompting their diagnosis. (Table 3: Early Development – Cardiac Concerns).

Among the 18 TS1 literature-only cases, prenatal cardiac concerns were discussed in 6 cases^10, 15–29^. Pre-term Caesarean births were performed due to bradycardia (5/6, 83%) or 2:1 AVB (3/6. 50%).

TS2 Gly406Arg surveys indicate 7/11 cases (64%) had cardiac presentations *in utero* (5 bradycardia and 4 2:1 AVB). Caesarean births were also common among pregnancies with TS2 Gly406Arg individuals: 9/11 (82%) of individuals were delivered by Caesarean section, and 4/9 of these caesarean births were pre-term (44%). One additional individual pre-term birth occurred via vaginal delivery. At birth, 9/11 (82%) TS2 infants were noted to have cardiac concerns (4 LQT, 8 bradycardia, and 7 with 2:1 AVB). (Table 3: Early Development – Cardiac Concerns).

In contrast, none of the Gly402Ser children presented *in utero* or at birth with any cardiac manifestations, nor were any delivered pre-term or by Cesarean section. None of the individuals with a diagnosis of nsLQT8 had cardiac concerns *in utero* or were born pre-term, but 3/12 were delivered by Caesarean section. For those with other variants in *CACNA1C*, 5/28 (18%) were identified to have cardiac concerns *in utero*. Five of 27 (19%) were born pre-term, and several were delivered by Caesarean section (9/27, 33%). (Table 3: Early Development – Cardiac Concerns).

#### Infancy to early childhood

Of the TS1 infants, when cardiac electrical concerns were noted, echocardiograms (echo) were performed near birth and often revealed congenital heart defects (CHDs) (11/30 cases, 37%), including patent ductus arteriosus (PDA), patent foramen ovale (PFO), ventricular septal defect (VSD), hypertrophic cardiomyopathy (HCM), and Tetralogy of Fallot (ToF). Eight respondents noted CHDs but were not specific in naming the defect. Thirteen of 29 survey respondents (45%) indicated that LQT was diagnosed at or near birth. (Table 3: Early Development – Cardiac Concerns).

Of the TS2 Gly406Arg children, 10/11 also had neonatal echo for detection of CHDs, and 2/10 (20%) children were identified as having one or more cardiac defects, such as PDA, VSD, and left ventricular hypertrophy (LVH) (Table 3: Early Development – Cardiac Concerns).

All Gly402Ser respondents appeared normal as infants with no apparent health concerns including no reported CHDs. The first cardiac presentation for all these children was sudden cardiac arrest (SCA) (average age 3.7 years) which they survived varying degrees of sequelae. Activities precipitating the Gly402Ser SCA events varied, ranging from physical activity, excitement, sitting quietly, or even sleeping. Upon cardiac evaluations obtained post SCA, all the children were noted to have a prolonged QT interval (average = 511.5ms, range 460-650ms). Of the six children whose families completed a survey, none reported apparent or recognizable signs of risk in the immediate family or extended family pedigree indicative of SUD, syncope, or cardiac concerns. Subsequent genetic testing suggested that 83% (5/6) of individuals acquired their variant *de novo*. (Table 3: Early Development – Cardiac Concerns).

In those diagnosed with nsLQT8, only 1/6 (17%) were identified to have an arrhythmia at birth, which was bradycardia. Two of 8 respondents (25%) diagnosed with nsLQT8 noted having a CHD, but were not specific in naming the defect. In the individuals with other *CACNA1C* variants, 13/23 (57%) were diagnosed with cardiac electrical concerns at birth. 11/24 were identified as having CHDs, including ToF, PDA, PFO, atrial septal defect, VSD, and HCM (Table 3: Early Development – Cardiac Concerns).

### Systematic evaluation of symptoms in TS

#### Sensitivity to anesthesia

Sensitivity to anesthesia is a serious threat to all TS individuals. Many children have experienced cardiac arrythmias upon administration of anesthesia (19/28 TS1 and 7/9 TS2 Gly406Arg) for correction of syndactyly, hip dysplasia, or dental procedures to remove diseased teeth^28^. In at least 12 TS1 cases (12/30; 40%), anesthesia was associated with the induction of ventricular arrhythmias during surgery. One TS1 child died after anesthesia administration. It is unclear whether hypoglycemia plays a role in this sensitivity, given that fasting is required prior to most surgical procedures. Half of Gly402Ser and diagnosed nsLQT8 individuals reported abnormal responses to anesthesia (3/6 and 5/10 respectively). Anesthesia is a risk for individuals with other variants as well, as 4/13 (31%) of these respondents reported an abnormal response to anesthesia. (Table 3: Early Development – Cardiac Concerns)

#### Hypoglycemia

While episodes of severe hypoglycemia were noted in the original description of TS ^10^; its potential to cause serious adverse outcomes became apparent after the death of a hypoglycemic child who did not have an ICD-documented arrhythmic event. Survey results indicated that hypoglycemia is a common concern for TS1 children. Fourteen of 26 (54%) TS1 individuals were reported as having had at least one known episode of hypoglycemia at some time in life. Six of the 14 reported that their child was hypoglycemic at birth (46%); the other eight parents reported at least one hypoglycemic event during childhood. Five of these eight children had two or more hypoglycemic episodes. Hypoglycemia also affects TS2 Gly406Arg individuals. For these children, 3/11 (27%) experienced an episode of hypoglycemia at birth. One of these three had at least two additional hypoglycemic episodes during childhood. (Table 4: Hypoglycemia and Endocrine Dysfunction)

**Table 4:** Hypoglycemia and Endocrine Dysfunction:

|  | TS1<br>Gly406Arg<br>Yes/Total (%)<br>n=30 | TS2<br>Gly406Arg<br>Yes/Total (%)<br>n=11 | Gly402Ser<br>Yes/Total (%)<br>n=6 | Diagnosed<br>nsLQT8<br>Yes/Total (%)<br>n=12 | Other<br>Yes/Total (%)<br>n=28 |
| --- | --- | --- | --- | --- | --- |
| Pancreas |  |  |  |  |  |
| Hypoglycemia | 14/26 (54) | 3/11 (27) | 2/4 (50) | 4/11 (36) | 10/26 (38) |
| Hypoglycemic at birth | 6/14 (43) | 3/11 (27) | 0/6 (0) | 0/6 (0) | 3/3 (100) |
| Hypoglycemic Death | 2/7 (29) | 0/1 (0) | 0/6 (0) | 0/11 (0) | --- |
| Pituitary Gland |  |  |  |  |  |
| Small birth size | 12/30 (40) | 4/10 (40) | 0/6 (0) | 1/11 (9) | 4/5 (80) |
| Growth hormone<br>replacement | 2/29 (7) | 1/10 (10) | 0/6 (0) | --- | 0/6 (0) |
| Early puberty | 0/6 (0) | 2/5 (40) | --- | 0/11 (0) | 0/10 (0) |
| Thyroid |  |  |  |  |  |
| Overweight | 3/30 (10) | 1/9 (11) | 1/6 (17) | 1/11 (9) | 4/26 (15) |
| Underweight | 5/30 (17) | 6/9 (67) | 0/6 (0) | 2/11 (18) | 7/26 (27) |
| Adrenal/Pineal Glands |  |  |  |  |  |
| Problems Sleeping | 12/29 (41) | 6/10 (60) | 2/6 (33) | 4/11 (36) | 13/23 (57) |
| Nightmares | 6/20 (30) | 0/5 (0) | 1/6 (17) | 2/7 (29) | 5/18 (28) |

Hypoglycemia was noted in two Gly402Ser individuals (2/4; 50%), and 4 of 11 (36%) survey respondents with diagnosed nsLQT8 reported at least one hypoglycemia episode, highlighting the potential for non-cardiac symptoms to arise in this category of patients. Many respondents in the other variant class (10/26; 38%) were affected, with 3 of these individuals reporting hypoglycemia at birth. Overall, hypoglycemia should be considered a serious life-threatening condition for all individuals with variants in *CACNA1C*.

#### Endocrine concerns

Forty percent (12/30) of TS1 newborns were considered small for gestational age (SGA). Thirty-one percent (9/29) of TS1 children remained underweight into childhood, and two required growth hormone therapy early in childhood (Table 4: Hypoglycemia and Endocrine Dysfunction). Forty percent (4/10) of TS2 Gly406Arg infants were considered SGA with 50% (5/10) remaining underweight into childhood; one required growth hormone therapy.

Although most TS children were too young to determine if precocious puberty was of concern, 6/6 TS individuals have reached puberty within the average range (8-14 years of age), while two female TS2 Gly406Arg children are being evaluated for precocious puberty. This was not a problem reported by any respondents diagnosed with nsLQT8 (0/11) or other respondents (0/10). Sleep disturbances were noted in 12/29 (41%) TS1, 6/10 (60%) TS2 Gly406Arg, and 2/6 (33%) Gly402Ser children. Problems sleeping were shared by those diagnosed with nsLQT8 and the other variant respondents, affecting 4/11 (36%) and 13/23 (57%) respectively. (Table 4: Hypoglycemia and Endocrine Dysfunction).

#### Pulmonary/respiratory concerns

Pulmonary dysfunction has been reported in TS children^10^. Survey data revealed pulmonary complications in TS1 and TS2 Gly406Arg children at birth and during infancy, with 10/24 (42%) of TS1 children experiencing pulmonary problems, some associated with PDA. One TS2 Gly406Arg child without PDA had primary pulmonary hypertension with right-to- left atrial level shunting and died due to hypoxic arrest (despite pulmonary vasodilator medications) without documentation of arrhythmia. Pulmonary presentations persisted throughout childhood in TS1 with 10/21 (48%) respondents reporting frequent pneumonias. Interestingly, 8/10 TS1 cases who reported frequent pneumonias were male. Other pulmonary concerns in the TS1 children included asthma and the need for nebulizer treatments for colds and infections. Three of 6 (50%) TS2 Gly406Arg children also suffered frequent pneumonias. However, respiratory concerns were not commonly reported in Gly402Ser individuals. (Table 5: Respiratory and Immune Dysfunction)

**Table 5:** Respiratory and Immune Dysfunction:

|  | TS1<br>Gly406Arg<br>Yes/Total (%)<br>n=30 | TS2<br>Gly406Arg<br>Yes/Total (%)<br>n=11 | Gly402Ser<br>Yes/Total (%)<br>n=6 | Diagnosed<br>nsLQT8<br>Yes/Total (%)<br>n=12 | Other<br>Yes/Total (%)<br>n=28 |
| --- | --- | --- | --- | --- | --- |
| Early childhood respiratory concerns: | 10/24 (42) | 2/10 (20) | 0/6 (0) | 3/4 (75) | 8/10 (80) |
| Aspiration | 2/29 (7) | 1/10 (10) | 1/6 (17) | 0/4 (0) | 4/10 (40) |
| Frequent pneumonia | 10/21 (48) | 3/6 (50) | 1/6 (17) | 0/4 (0) | 6/10 (60) |
| Asthma | 7/23 (30) | 1/9 (11) | 0/6 (0) | 3/4 (75) | 1/10 (10) |
| Nebulizer treatments | 5/20 (25) | 1/9 (11) | 0/6 (0) | 3/4 (75) | 0/10 (0) |
| Adult respiratory concerns: |  |  |  |  |  |
| CRAP use | 1/6 (17) | --- | --- | --- | --- |
| Respiratory deaths | 1/7 (14) | 1/2 (50) | --- | --- | 2/3 (67) |
| Frequent infections | 9/25 (36) | 5/11 (45) | 0/6 (0) | 9/11 (82) | 11/21 (52) |
| Delayed wound healing | 2/30 (6) | 1/9 (11) | 0/6 (0) | 2/9 (22) | 4/19 (23) |

Three out of 4 (75%) of those diagnosed with nsLQT8 reported respiratory concerns in early childhood. Survey respondents with other variants noted respiratory concerns in early childhood (8/10, 80%), most commonly frequent pneumonia (6/10, 60%). Additionally, two individuals died from respiratory arrest: one from respiratory syncytial virus requiring O_2_ at time of death and one from respiratory failure. An additional individual had a chest infection at death requiring an x-ray and suffered cardiac arrest during the x-ray. This suggests that respiratory concerns and infection are common and potentially life-threatening among individuals harboring a *CACNA1C* variant.

#### Immune system

Thirty-six percent (9/25) of TS1 respondents reported frequent, recurring infections. One TS1 individual was found to have IgG2 deficiency, presumed to be responsible for infectious complications of bronchopneumonia, bronchitis, obstructive lung disease and asthma. Forty-five percent (5/11) of TS2 Gly406Arg children reported having frequent infections, though the exact nature of these infections was unknown. While no Gly402Ser individuals had immunological concerns, those with other variants in *CACNA1C* reported frequent infections, including 9/11 (82%) respondents diagnosed with nsLQT8 and 11/21 (51%) of those within the other variant category. The extent of immune dysfunction in TS individuals is otherwise not well understood given that most have not undergone comprehensive testing for immune deficiencies, however it does appear to be a recurring feature across the patient population. (Table 5: Respiratory and Immune Dysfunction).

#### Syndactyly

Syndactyly in combination with arrythmias are the most common characteristics of the TS1 infant^10^ and are often recognized at birth. The syndactyly observed in TS1 newborns has been classified as simple (skin and soft tissue only) or complex (skin, soft tissue, and bone). Ninety-six percent of TS1 cases (45/47, respondents and literature cases combined) had either hand and/or foot syndactyly, with at least 89% (42/47) of genotyped positive TS1 children having obvious syndactyly (one or both hands). Survey results of TS1 individuals indicated 26/30 (87%) had obvious hand syndactyly, while two additional children, upon closer examination, exhibited slight simple syndactyly, totaling 28/30 (93%) in our cohort (Table 6: Skeletal, Facial, Skin, and Dental).

**Table 6:** Skeletal, Facial, Skin, and Dental.

|  | TS1<br>Gly406Arg<br>Yes/Total (%)<br>n=30 | TS2<br>Gly406Arg<br>Yes/Total (%)<br>n=11 | Gly402Ser<br>Yes/Total (%)<br>n=6 | Diagnosed<br>nsLQT8<br>Yes/Total (%)<br>n=12 | Other<br>Yes/Total (%)<br>n=28 |
| --- | --- | --- | --- | --- | --- |
| Bone development abnormalities: |  |  |  |  |  |
| Finger Syndactyly | 28/30 (93) | 1/11 (9) | 0/6 (0) | 0/11 (0) | 13/27 (48) |
| Includes 4-5 | 26/28 (93) | 1/11 (9) | --- | --- | 10/27 (37) |
| Bilateral | 18/28 (64) | 1/11 (9) | --- | --- | 7/27 (26) |
| Toe Syndactyly | 24/30 (80) | 1/11 (9) | 0/6 (0) | 0/10 (0) | 11/27 (41) |
| 2-3 toe | 23/24 (96) | 1/11 (9) | --- | --- | 10/27 (37) |
| Bilateral | 23/24 (96) | 1/11 (9) | --- | --- | 10/27 (27) |
| Hip Dysplasia | 1/30 (3) | 3/9 (33) | 0/6 (0) | 0/3 (0) | 2/17 (12) |
| Joint contractors | 0/30 (0) | 6/8 (75) | 0/6 (0) | 0/3 (0) | 0/17 (0) |
| Club foot | 0/30 (0) | 3/11 (27) | 0/6 (0) | 0/3 (0) | 1/17 (6) |
| Facial abnormalities |  |  |  |  |  |
| Round face | 23/25 (92) | 7/9 (78) | 3/6 (50) | 2/3 (67) | 12/21 (57) |
| Broad forehead | 23/23 (100) | 7/9 (78) | 4/6 (67) | --- | 5/21 (29) |
| Thin upper lip | 22/22 (100) | 7/9 (78) | 6/6 (100) | 0/3 (0) | 2/21 (13) |
| Depressed maxillary | 5/23 (22) | 0/7 (0) | 0/6 (0) | 0/3 (0) | 0/21 (0) |
| Low set ears | 11/22 (50) | 7/9 (78) | 0/6 (0) | 1/3 (33) | 8/21 (38) |
| Flat bridge nose | 6/15 (40) | 2/4 (50) | 1/6 (17) | 0/3 (0) | 7/21 (33) |
| Hypertelorism | 10/22 (45) | --- | 0/6 (0) | --- | 1/21 (5) |
| Flat philtrum | 6/21 (29) | 1/9 (11) | 0/6 (0) | 0/3 (0) | 0/21 (0) |
| Bald at birth | 26/27 (96) | 2/11 (18) | 1/6 (17) | 1/3 (33) | 11/21 (52) |
| Skin abnormalities: |  |  |  |  |  |
| Skin sloughing | 7/20 (35) | 2/2 (100) | 0/6 (0) | 1/7 (14) | 5/12 (42) |
| Odd color | 9/20 (45) | 1/5 (20) | 1/6 (17) | 2/7 (29) | 1/12 (8) |
| Hot/cold sensitivity | 26/27 (96) | 2/11 (18) | 3/6 (50) | 5/7 (71) | 9/12 (75) |
| Dental abnormalities |  |  |  |  |  |
| Frequent cavities | 9/22 (41) | 3/5 (60) | 0/6 (0) | 3/9 (33) | 7/21 (33) |
| Small teeth | 11/18 (61) | 2/5 (40) | 0/6 (0) | 2/9 (22) | 3/21 (14) |
| Misplaced teeth | 9/11 (82) | 3/3 (100) | 1/6 (17) | 3/9 (33) | 6/21 (29) |
| Tooth extraction surgery | 7/16 (44) | 4/7 (57) | 1/6 (17) | 4/9 (44) | 6/21 (29) |
| Sedation concerns | 4/9 (44) | 2/2 (100) | --- | 0/9 (0) | 1/21 (5) |

Syndactyly of 4th and 5^th^ digit among TS1 children, an atypical presentation that differs from the typical fusion of the third and fourth finger, was noted in a recent study that included children from our cohort^33^. Our survey results identified that 87% (26/28) of TS1 children with obvious syndactyly presented with inclusion of the 4th and 5^th^ digit, with the most frequent finger syndactyly including digits 3-5 (n=15), 4-5 (n=8) and 2-5 (n=3). Of the 26 children with obvious syndactyly that included the 5^th^ digit, 16/26 (62%) had bilateral involvement, with four (15%) having only left-hand 4-5 syndactyly and six (23%) having different finger involvement in each hand. The remaining two of 30 (7%) TS1 individuals have classic bilateral 3-4 syndactyly. (Table 6: Skeletal, Facial, Skin, and Dental).

Unlike TS1, TS2 has not been associated with syndactyly, allowing for phenotypic distinction between the two types of TS^10, 13^. Only one TS2 Gly406Arg child was noted as having syndactyly; however, this child has a secondary genetic change [4q31.3q32.1(155,189,952-155,654,941)x1], which is sporadically associated with 5^th^ digit abnormalities and webbing between fingers^34^. Likewise, no respondents with Gly402Ser or diagnosed with LQT8 reported any syndactyly of fingers or toes. Unfortunately, this lack of a recognizable physical anomaly may reduce identification of these infants at birth. (Table 6: Skeletal, Facial, Skin, and Dental).

Survey respondents in the other variant class fall in the middle, with 13/27 (48%) reporting finger syndactyly and 11/27 (41%) reporting toe syndactyly. Ten of the 13 with finger syndactyly have fusion of digits 4-5, matching the TS1 hallmark.

#### Bones and joints

TS2 Gly406Arg individuals are distinguished from TS1 by the presence of skeletal development abnormalities, namely hip dysplasia, which we identified in 3/9 (33%) TS2 Gly406Arg respondents. TS2 Gly406Arg children are also identified as having joint contractures of the hands and feet (6/8; 75%) and/or a club foot (3/11; 27%). Radial head dysplasia was noted in a pair of TS2 Gly406Arg twins and protrusion of the distal end of the sternum was noted in two children. Six TS2 Gly406Arg respondents also noted joint hypermobility in an open-ended request for additional concerns. These abnormalities were generally absent in TS1 individuals. Twenty-nine of 30 (97%) TS1 children lacked any bone abnormalities noted in TS2 Gly406Arg children (other than syndactyly), however, one TS1 child was noted to have hip dysplasia. No bone and joint concerns were reported by Gly402Ser or diagnosed nsLQT8 individuals. Bone and joint concerns were not common in patients harboring other variants, with just two individuals reporting hip dysplasia (2/17; 12%) and one with club foot (1/17; 5%). (Table 6: Skeletal, Facial, Skin, and Dental).

#### Hair growth, macular-facial development

A round face is a common feature of TS1 children (92%, 23/25) and this facial shape generally remains into adult life (5 of 6 TS1 surviving adults have retained the appearance of a round, infantile face). Round faces are also common in TS2 children (78%, 7/9). A broad forehead or a receding hairline is noted in all TS1 children and in 78% (7/9) of TS2 Gly406Arg children. Another hallmark facial feature noted in 22/22 (100%) TS1 children and 7/9 (78%) TS2 Gly406Arg children is a thin upper lip. In TS1, 6/21 (29%) have a flat philtrum, particularly recognized when smiling; this appears to be an infrequent characteristic in TS2 Gly406Arg individuals (1/9, 11%). A depressed maxilla or a hypertrophied mandible is noted in 5/23 (22%) TS1 children, but this facial feature was not recognized in any (0/7) of TS2 Gly406Arg children. Low set ears are noted in 11/22 (50%) TS1 and 7/9 (78%) TS2 Gly406Arg children. A flat nasal bridge was reported in 6 of 15 (40%) TS1 and 2 of 4 (50%) TS2 Gly406Arg children. (Table 6: Skeletal, Facial, Skin, and Dental).

Gly402Ser individuals also had several of these features: three had round faces (50%), four had broad foreheads (67%), all six had thin upper lips, one had low set ears (17%), but none had a flat philtrum or depressed maxillary. Two diagnosed nsLQT8 patients reported facial abnormalities, however only 3 of the 12 survey respondents answered this question, limiting conclusions. Among the other variant respondents, a round face and low set ears were most commonly reported (12/21; 57% and 8/21; 38%, respectively). Thus, facial abnormalities appear to be common among individuals with *CACNA1C* variants, with some differences in pattern across the different categories.

Ninety-six percent (26/27) of TS1 newborns were born bald or with sparse hair. Only 18% (2/11) of TS2 Gly406Arg newborns had little or no hair at birth. The Gly402Ser respondents were similar to the TS2 Gly406Arg group, with only 1 of 6 (17%) reporting little or no hair at birth. Other cases fell in the middle, with 33% (1/3) of those diagnosed with nsLQT8 and 52% (11/21) of other patients reporting baldness at birth.

#### Skin – dermatologic and sensory abnormalities

Skin disorders were noted among individuals with *CACNA1C* variants across the different groups. No respondents reported having the autoimmune disease of psoriasis, however skin sloughing, unrelated to environmental or geographical living conditions, was reported by 7/20 (35%) TS1, 2/2 TS2 Gly406Arg, 1/7 (14%) diagnosed nsLQT8 and 5/12 (42%) other respondents. Odd, strange, or diagnostically undetermined skin color anomalies were reported by 9/20 (45%) TS1, 1/5 (20%) TS2 Gly406Arg, 1/6 (17%) Gly204Ser, 2/7 (29%) diagnosed nsLQT8, and 1/12 (8%) other respondents. (Table 6: Skeletal, Facial, Skin, and Dental).

#### Dental features

Forty-one percent (9/22) of TS1 children had frequent, large dental caries/decay, with 7 of 16 (44%) requiring surgical removal of multiple diseased teeth. Three of 5 TS2 Gly406Arg children also struggled with frequent cavities (60%). Small teeth were noted in 11 of 18 (61%) TS1 and 2 of 5 (40%) TS2 Gly406Arg children. Misplaced teeth were reported in 9/11 (82%) TS1 respondents and 3/3 TS2 Gly406Arg children. Individuals with Gly402Ser did not report frequent cavities or small teeth, but 1/6 (17%) reported misplaced teeth and subsequent corrective surgery. A few diagnosed nsLQT8 also reported dental concerns, including frequent cavities (3/9; 33%), misplaced teeth (3/9; 33%), and tooth extraction surgery (4/9; 44%). Other *CACNA1C* variant individuals reported frequent cavities (7/21; 33%), misplaced teeth (6/21; 29%), and tooth extraction surgery (6/21; 29%). (Table 6: Skeletal, Facial, Skin, and Dental)

#### Skeletal muscle

Hypotonia is present in both TS1 (5/26, 19%) and TS2 Gly406Arg (5/7, 71%), but appears to be more frequent and severe in TS2 Gly406Arg, as evidenced by delay or impairment in walking. Only one Gly402Ser individual reported hypotonia (1/6; 17%). Although walking unassisted was delayed in some TS1 children, all children have learned to walk. Among survey respondents, all parents of TS2 Gly406Arg children considered their children to be lacking in physical coordination. Of note, one child requires mechanical assistance to walk, and two children are non-ambulatory. As TS2 Gly406Arg children transition into older childhood, other muscular concerns become apparent. Many patients experience disproportionate upper versus lower body development (4/7; 57%) and body strength discrepancies between right and left side (3/5; 60%). In contrast, these characteristics were not reported by any TS1individuals. However, coordination issues were reported in TS1 (11/13; 61%), TS2 Gly406Arg (5/5, 100%), Gly402Ser children (2/6; 33%), diagnosed nsLQT8 (4/7; 57%), and other variant cases (11/19; 58%) (Table 7: Neuromuscular and Sensory Concerns).

**Table 7:** Neuromuscular and Sensory Concerns:

|  | TS1<br>Gly406Arg<br>Yes/Total (%)<br>n=30 | TS2<br>Gly406Arg<br>Yes/Total (%)<br>n=11 | Gly402Ser<br>Yes/Total (%)<br>n=6 | Diagnosed<br>nsLQT8<br>Yes/Total (%)<br>n=12 | Other<br>Yes/Total (%)<br>n=28 |
| --- | --- | --- | --- | --- | --- |
| Neuro-muscular concerns |  |  |  |  |  |
| Hypotonia | 5/26 (19) | 5/7 (71) | 1/6 (17) | 1/3 (33) | 11/18 (61) |
| Severe weakness | 3/22 (14) | 3/7 (43) | 1/6 (17) | --- | 6/20 (30) |
| Strength differences | 0/28 (0) | 3/5 (60) | 0/6 (0) | --- | --- |
| Uneven body growth | 0/30 (0) | 4/7 (57) | 0/6 (0) | --- | 0/1 (0) |
| Lack of coordination | 11/13 (61) | 5/5 (100) | 2/6 (33) | 4/7 (57) | 11/19 (58) |
| Delayed milestones |  |  |  |  |  |
| Holding head (>3mos) | 11/18 (61) | 3/3 (100) | 1/3 (33) | 5/9 (56) | 11/20 (55) |
| Sitting (>6mos) | 15/19 (79) | 4/5 (80) | 0/4 (0) | 2/9 (22) | 12/17 (71) |
| Crawling (>9mos) | 14/18 (78) | 4/4 (100) | 0/3 (0) | 1/9 (11) | 12/14 (86) |
| Standing (>12mos) | 15/20 (75) | 7/7 (100) | 1/5 (20) | 1/9 (11) | 9/15 (60) |
| Walking (>18mos) | 10/20 (50) | 5/6 (83) | 5/5 (100) | 1/9 (11) | 8/17 (47) |
| Running (>24mos) | 8/18 (44) | 4/5 (80) | 5/5 (100) | 1/9 (11) | 6/15 (40) |
| Climbing (>24mos) | 8/16 (44) | 5/6 (83) | 5/5 (100) | 2/8 (25) | 8/14 (57) |
| Jumping (>30mos) | 12/16 (67) | 5/5 (100) | 1/4 (25) | 2/9 (22) | 8/12 (67) |
| Neurosensory concerns |  |  |  |  |  |
| Diagnosed eye conditions |  |  |  |  |  |
| Wears glasses | 14/24 (58) | 6/7 (86) | 2/6 (33) | 7/10 (70) | 11/23 (48) |
| Myopia | 3/14 (21) | 1/6 (17) | 0/2 (0) | 1/4 (25) | 0/17 (0) |
| Hyperopia | 2/14 (14) | 0/6 (0) | 0/2 (0) | --- | 1/17 (6) |
| Amblyopia | 1/14 (7) | 3/6 (50) | 0/2 (0) | 1/4 (25) | 6/17 (35) |
| Strabismus | 3/14 (21) | 2/6 (33) | 1/2 (50) | 0/4 (0) | 2/17 (12) |
| Astigmatism | 3/14 (21) | 0/6 (0) | 0/2 (0) | 4/4 (100) | 6/17 (35) |
| Retina Concern | 2/14 (14) | 0/6 (0) | 0/2 (0) | 0/4 (0) | 0/17 (0) |
| Cortical Blindness | 0/30 (0) | 1/7 (14) | 0/6 (0) | 0/4 (0) | 1/17 (6) |
| Diagnosed hearing concern | 0/30 (0) | 0/11 (0) | 0/6 (0) | 3/8 (38) | 1/20 (5) |
| Sounds irritate | 11/25 (44) | 5/10 (50) | 2/6 (33) | 7/11 (64) | 13/21 (62) |
| Taste Concerns |  |  |  |  |  |
| Food aversion | 2/25 (8) | --- | 2/6 (33) | --- | 2/21 (10) |
| Food textures | 6/25 (19) | 2/8 (25) | 2/6 (33) | 3/7 (43) | 8/21 (38) |
| Olfactory Concern | 0/30 (0) | 0/9 (0) | 1/6 (17) | --- | 0/1 (0) |
| Tactile Concern |  |  |  |  |  |
| Clothing preferences | 8/20 (40) | 3/6 (50) | 2/6 (83) | 5/9 (56) | 10/20 (50) |

#### Neurosensory

Many TS children have vision abnormalities requiring corrective lenses. Fifty-eight percent (14/24) of TS1 children wear corrective glasses for vision concerns including myopia (3/14), astigmatism (3/14), amblyopia (1/14), hyperopia (2/14), retina concerns (2/14), strabismus (3/14), and coloboma of the iris (1/14). Of 11 TS2 Gly406Arg respondents, 7 have had vision examinations. At the time of the survey, six required corrective lenses; 2 for strabismus, 3 for amblyopia, and 1 for myopia, while the remaining child was found to be cortically blind. Only 2/6 (33%) of Gly402S respondents had eye concerns, while glasses were commonly reported in LQT8 individuals (7/10; 70%). However, this higher percentage could reflect the older age of this cohort. Forty-eight percent (11/23) of other *CACNA1C* variant individuals required glasses, with the most common visual concerns being amblyopia (6/17; 35%) and astigmatism (6/17; 35%). (Table 7: Neuromuscular and Sensory Concerns)

All TS1 and TS2 Gly406Arg and Gly402Ser children appeared to have normal or near normal hearing. The trend was similar in other variant individuals with only 1 individual reporting a diagnosed hearing concern (1/20; 5%). Hearing loss was most common in individuals diagnosed with nsLQT8 (3/8; 38%), though this could again reflect the older age of the cohort. (Table 7: Neuromuscular and Sensory Concerns)

Sounds and particularly loud noises were reported as irritating and causing distress in 11 of 25 (44%) TS1 children, 5/10 (50%) TS2 children, and 2/6 (33%) Gly402S respondents. This was similar in the other variant group, with 62% (13/21) reporting irritation from loud noises (Table 7: Neuromuscular and Sensory Concerns).

#### Speech concerns

Sixty-nine percent (18/26) of TS1 parents surveyed considered their children to be speech delayed, and 6/9 (67%) TS2 Gly406Arg parents reported the same concern. Of the six TS2 Gly406Arg children considered delayed in speech, two are non-verbal, though one is adept at communicating with sign language and the written word on a laptop computer. Ten of 20 (50%) TS1 and 5/7 (71%) TS2 Gly406Arg children showed a delay in receptive speech understanding. Greater delays in expressive speech were reported for 13 of 20 (65%) TS1 and 5 of 7 (71%) TS2 Gly406Arg children. Twenty percent (5/25) of TS1 children expressed themselves with odd sounds or had articulation disorders; 25% (2/8) of TS2 Gly406Arg children also exhibited similar articulation disorders. Speaking in full sentences was delayed in both TS1 and TS2 Gly406Arg individuals, as reported by parents. The average age at which TS1 children spoke in full sentences was 4.75 years (19/26). Only two TS2 children reported an age at which they spoke in full sentences, which averaged 4.75 years. Two out of 6 (33%) Gly402Ser respondents reported speech delay, which was similar for patients diagnosed with nsLQT8 (3/10; 30%). In contrast, speech delay was more common in other variant individuals (15/17; 88%). (Table 8: Neuropsychological Characteristics and Adult Life Skills)

**Table 8:** Neuropsychological Characteristics and Adult Life Skills:

|  | TS1<br>Gly406Arg<br>Yes/Total (%)<br>n=30 | TS2<br>Gly406Arg<br>Yes/Total (%)<br>n=11 | Gly402Ser<br>Yes/Total (%)<br>n=6 | Diagnosed<br>nsLQT8<br>Yes/Total (%)<br>n=12 | Other<br>Yes/Total (%)<br>n=28 |
| --- | --- | --- | --- | --- | --- |
| Neuropsychological concerns |  |  |  |  |  |
| Speech |  |  |  |  |  |
| Speech delay | 18/26 (69) | 6/9 (67) | 2/6 (33) | 3/10 (30) | 15/17 (88) |
| Receptive delay | 10/20 (50) | 5/7 (71) | 2/6 (33) | --- | 5/17 (29) |
| Expressive delay | 13/20 (65) | 5/7 (71) | 2/6 (33) | 2/4 (50) | 12/17 (71) |
| Odd sounds | 5/25 (20) | 2/8 (25) | 2/6 (33) | 1/4 (25) | 9/17 (53) |
| Educational |  |  |  |  |  |
| Global developmental delay | 0/30 (0) | 2/10 (20) | 0/6 (0) | --- | 4/5 (80) |
| Knows alphabet | 13/17 (76) | 1/4 (25) | 6/6 (100) | 12/12 (100) | 9/20 (45) |
| At grade level | 5/17 (29) | 0/5 (0) | 0/1 (0) | 9/11 (82) | 3/15 (20) |
| Problems reading | 12/16 (75) | 1/4 (25) | 3/6 (50) | 2/9 (22) | 12/15 (80) |
| Problems with numbers | 10/15 (67) | 4/4 (100) | 3/6 (50) | 4/9 (44) | 10/15 (67) |
| Accurate memory | 8/18 (44) | 1/4 (25) | 1/4 (25) | 2/9 (22) | 2/15 (13) |
| Delayed potty training (>4yrs) | 10/20 (50) | 7/8 (86) | 2/4 (50) | 2/7 (29) | 2/7 (29) |
| Delayed night training (>5yrs) | 7/17 (41) | 7/7 (100) | 1/4 (25) | 4/10 (40) | --- |
| Social ability |  |  |  |  |  |
| Not tested | 13/19 (68) | 1/5 (20) | 3/6 (22) | --- | --- |
| Autism | 6/12 (50) | 4/9 (44) | 1/3 (33) | 2/8 (25) | 8/14 (57) |
| ADD/ADHD | 5/11 (45) | 5/9 (56) | 3/5 (60) | 1/8 (13) | 4/14 (29) |
| Shy/anxious | 14/24 (58) | 4/7 (57) | 5/6 (83) | 2/10 (2) | 7/19 (37) |
| Odd behaviors | 13/23 (57) | 4/5 (80) | 1/4 (25) | 2/10 (2) | 6/19 (32) |
| Plays with age group | 12/23 (52) | 2/4 (50) | 1/4 (25) | 6/10 (60) | 8/19 (42) |
| Prefers being alone | 11/21 (52) | 2/4 (50) | 2/6 (33) | 5/10 (50) | 6/19 (32) |
| Prefers quiet environment | 14/21 (67) | 3/5 (60) | 2/6 (33) | 3/10 (30) | 7/19 (37) |
| Prefers animals | 8/19 (42) | 1/5 (25) | 0/6 (0) | 3/10 (30) | 4/19 (21) |
| Other neural development |  |  |  |  |  |
| Routine oriented | 17/22 (77) | 5/5 (100) | 4/6 (67) | 5/10 (50) | 9/19 (47) |
| Orders objects | 9/23 (39) | 1/3 (33) | 1/4 (25) |  | 6/19 (32) |
| Seizure disorder | 10/30 (33) | 5/7 (71) | 0/6 (0) | 1/11 (9) | 11/22 (50) |
| Has phobias | 9/24 (38) | 0/4 (0) | 0/6 (0) | 2/8 (25) | 1/14 (7) |
| Anger/violence | 11/23 (48) | 2/5 (40) | 2/6 (33) | 1/8 (13) | 4/14 (29) |
| Depression | 1/6 (17) | --- | 0/6 (0) | 2/8 (25) | 2/14 (14) |
| OCD | 4/19 (21) | --- | 1/6 (17) | 2/8 (25) | 3/14 (21) |
| Schizophrenia | 1/19 (5) | --- | 1/6 (17) | 0/8 (0) | 0/14 (0) |
| Severe headaches | 2/10 (11) | 0/2 (0) | 2/6 (33) | 1/8 (13) | 1/14 (7) |
| Adult life skills |  |  |  |  |  |
| Rides bus alone | 2/6 (33) | --- | --- | 6/6 (100) | --- |
| Has driver's license | 1/6 (17) | --- | --- | 6/6 (100) | --- |
| Makes monetary change | 1/6 (17) | --- | --- | 6/6 (100) | --- |

#### Neurodevelopment and educational/learning delays

The TS1 and TS2 Gly406Arg school age population is small, but most experience educational and learning delays. Thirteen of 17 (76%) of the TS1 children were able to learn the alphabet. Twelve of 16 (75%) TS1 and 1 of 4 (25%) TS2 Gly406Arg children have not attained age-appropriate reading level skills, and 10 of 15 (67%) TS1 and 4 of 4 TS2 Gly406Arg children lack numerical/computational skills. Three of 6 (50%) Gly402Ser children have problems with reading and math, although these delays are difficult to separate from potential effects of SCA events. Problems with reading and math skills were less prevalent in diagnosed nsLQT8 individuals (2/9; 22% and 4/9 22%, respectively), but common in individuals with other variants (12/15; 80% and 10/15; 67%, respectively). Delayed toilet training (>4 years old) is noted in 10/20 (50%) of TS1 and 7/8 (86%) of TS2 Gly406Arg children. Nighttime training was delayed in TS1 (7/17, 41%) and TS2 Gly406Arg (7/7, 100%) children. (Table 8: Neuropsychological Characteristics)

Interestingly, 8/18 (44%) TS1 and 1/4 (25%) TS2 Gly406Arg children were described as having an “amazing and accurate memory” by parents. 6 of 12 (50%) TS1 survey respondents reported a diagnosis of autism, however this number is lower than earlier reports indicating as many as 80% of these patients suffer from Autism Spectrum Disorder ^10^. Thus, it is possible that the relatively low number of respondents answering this question and the young age of many of the individuals contributed to the lower value. In addition, 5/11 (45%) TS1 respondents reported attention-deficit disorder (ADD) or attention-deficit/hyperactive disorder (ADHD). Of nine TS2 Gly406Arg respondents, all were diagnosed with either autism (4/9) or ADD/ADHD (5/9) (Table 8: Neuropsychological Characteristics).

Seizure disorders were noted in 10/30 (33%) TS1 and 5/7 (71%) TS2 Gly406Arg individuals. Concerns of depression were seldom noted in TS1 (1/6, 17%), and were not reported in any TS2 Gly406Arg survey. Fifty-eight percent (14/24) of TS1 children were noted to be shy/anxious by their parents, as were 57% (4/7) of TS2 Gly406Arg children. Thirty-eight percent (9/24) of TS1 children were noted to have phobias, while this was not noted in TS2 Gly406Arg. Episodes of anger/violence were often noted in TS1 (11/23, 48%) and TS2 Gly406Arg (2/5, 40%) children. One TS1 individual has been diagnosed with schizophrenia (Table 8: Neuropsychological Characteristics).

A few Gly402Ser children report psychosocial concerns, including 3/5 (60%) with diagnoses of ADD/ADHD and 1/3 (33%) diagnosed with autism. Each of these conditions was only noted in 1-2 individuals diagnosed with nsLQT8. Individuals with other variants reported autism and ADD/ADHD diagnoses (8/14; 57% and 4/14; 29%, respectively) and 50% (11/22) noted seizure disorders. (Table 8: Neuropsychological Characteristics).

#### Gastrointestinal and urinary systems

Gastrointestinal and urinary dysfunction have been noted as additional symptoms of TS as more individuals are living longer and are followed over time. Early in life, esophageal dysfunction manifested as dysphagia for 13/28 (46%) TS1 and 7/10 (70%) TS2 Gly406Arg children. Seven of 27 (26%) TS1 and 4/10 (40%) TS2 Gly406Arg respondents reported suffering from gastric reflux. Twelve of 29 (41%) TS1 and 6/10 (60%) TS2 Gly406Arg individuals reported severe gag reflex. Nasogastric tube use was reported for 9/25 (36%) TS1 and 5/10 (50%) TS2 Gly406Arg individuals. Frequent vomiting also occurred in 7/28 (25%) TS1 and 6/9 (67%) TS2 Gly406Arg individuals. Only one out of 6 (17%) Gly402Ser respondents noted any esophageal concerns, which were gastric reflux and frequent vomiting. Gastric reflux was common in diagnosed nsLQT8 and individuals with other variants, affecting 5/7 (71%) and 12/22 (55%) respectively. (Table 9: Gastrointestinal and Urinary Dysfunction).

**Table 9:** Gastrointestinal and Urinary Dysfunction:

|  | TS1<br>Gly406Arg<br>Yes/Total (%)<br>n=30 | TS2<br>Gly406Arg<br>Yes/Total (%)<br>n=11 | Gly402Ser<br>Yes/Total (%)<br>n=6 | Diagnosed<br>nsLQT8<br>Yes/Total (%)<br>n=12 | Other<br>Yes/Total (%)<br>n=28 |
| --- | --- | --- | --- | --- | --- |
| Esophageal concerns |  |  |  |  |  |
| Difficulty swallowing | 13/28 (46) | 7/10 (70) | 0/6 (0) | --- | 7/22 (32) |
| Gastric reflux | 7/27 (26) | 4/10 (40) | 1/6 (17) | 5/7 (71) | 12/22 (55) |
| Frequent gagging | 12/29 (41) | 6/10 (60) | 0/6 (0) | 2/7 (29) | 4/22 (18) |
| Nasogastric tube | 9/25 (36) | 5/10 (50) | 0/6 (0) | 1/8 (13) | 9/22 (41) |
| Frequent vomiting | 7/28 (25) | 6/9 (67) | 1/6 (17) | 2/7 (29) | 6/22 (27) |
| Surgical procedures | 2/30 (7) | --- | --- | --- | --- |
| Stomach concerns |  |  |  |  |  |
| Frequent stomach aches | 9/27 (33) | 1/3 (33) | 2/4 (50) | 3/7 (43) | 12/25 (48) |
| Omphalocele | 0/30 (0) | 1/11 (9) | 0/6 (0) | --- | 0/5 (0) |
| Ileal atresia/dilation | 1/30 (3) | 1/5 (20) | 0/6 (0) | --- | 1/5 (20) |
| Elimination concerns |  |  |  |  |  |
| Constipation | 11/19 (38) | 6/10 (60) | 5/6 (83) | 7/7 (100) | 21/25 (90) |
| Diarrhea | 2/28 (7) | 2/10 (20) | 0/4 (0) | 4/7 (57) | 4/25 (16) |
| Urinary Concerns |  |  |  |  |  |
| Kidney abnormalities | 3/10 (30) | 0/11 (0) | 0/6 (0) | 1/10 (10) | 1/19 (5) |
| Body swelling | 2/10 (20) | 0/11 (0) | 0/6 (0) | --- | 0/5 (0) |

Severe stomach aches were noted in 33% of TS1 (9/27) and TS2 Gly406Arg cases (1/3). Half of Gly402Ser respondents (2/4) and nearly half of those diagnosed with nsLQT8 (3/7; 43%) or other variants (12/25; 48%) also reported frequent severe stomach aches. Anatomic gastrointestinal defects that were reported included ileal atresia/dilation (1/ 30, 3% of TS1 children) and omphalocele with bowel malrotation (1/11 (9%) TS2 Gly406Arg children). These abnormalities required surgical repair.

Diarrhea has not been a major concern, affecting 2/28 children with TS1 (75) and 2/10 TS2 Gly406Arg children (20%). Constipation was more common across all groups, reported in 11 of 19 (38%) TS1 and 6 of 10 (60%) TS2 Gly406Arg children. Two TS1 children reported receiving emergency room attention due to sustained penile pain and tumescence resulting from severe fecal blockage within the bowel. Penile pain is considered common in the pediatric population, resulting from several etiologies^35^. Severe constipation can exert external pressure on the bladder wall, causing referred pain to the genitalia. In the two TS1 children who experienced this, emergency room assistance was required to remove the bowel blockage and relieve the pain. Constipation was common across TS groups, affecting 5/6 (83%) of Gly402Ser respondents, 7/7 (100%) diagnosed nsLQT and 21/25 (90%) other variant respondents.

As the TS1 population ages, there is growing evidence of kidney dysfunction (3/10; 30%). One adult woman was diagnosed with stage three kidney failure, ascribed to scarring associated with frequent urinary tract infections and severe constipation.

### Current Therapies

#### Implantable Cardiac Defibrillators (ICDs)

Most of the surviving TS1 (24/30, 80%), TS2 Gly406Arg (9/11, 82%), Gly402Ser (6/6, 100%), and other *CACNA1C* variant (6/16, 38%) children reported having an ICD implanted early in life or soon after a cardiac event. Six TS1 individuals currently do not have ICDs due to their small size or parental decision; each family reported having an AED with them at all times. Currently two TS2 Gly406Arg children are managed with AEDs only. Of the 8/30 TS1 individuals who have died, 63% (5/8) were too young to consider ICD implantation. Although the other 3/8 had ICDs at the time of their deaths, ICD interrogation revealed no arrhythmias in two of the three. Of the three deaths among individuals with TS2 Gly406Arg (11 total), only one had an ICD at the time of death and interrogation revealed no precipitant arrhythmia. The causes of death of the other two children with TS2 Gly406Arg remain unknown. Two individuals with Gly402Ser have suffered multiple ICD shocks that have continued despite left cardiac stellate denervation (LCSD) surgery (see below). (Table 10: Cardiac Medications and Procedures)

**Table 10:** Cardiac Medications and Procedures:

|  | TS1<br>Gly406Arg<br>Yes/Total (%)<br>n=30 | TS2<br>Gly406Arg<br>Yes/Total (%)<br>n=11 | Gly402Ser<br>Yes/Total (%)<br>n=6 | Diagnosed<br>nsLQT8<br>Yes/Total (%)<br>n=12 | Other<br>Yes/Total (%)<br>n=28 |
| --- | --- | --- | --- | --- | --- |
| Implanted Devices |  |  |  |  |  |
| Infant Pacemaker | 9/30 (30) | 5/11 (45) | 0/6 (0) | 0/12 (0) | 1/21 (10) |
| Infant ICD | 7/30 (23) | 1/11 (9) | 0/6 (0) | 0/12 (0) | 0/26 (0) |
| Other ages ICD | 17/30 (57) | 8/8 (100) | 6/6 (100) | --- | 6/16 (38) |
| Infant LCSD | 3/30 (10) | 1/11 (9) | 0/6 (0) | --- | 1/5 (20) |
| Other ages LCSD | 4/30 (13) | 0/10 (0) | 2/6 (33) | --- | 0/4 (0) |
| Beta-Blockers | 27/30 (93) | 11/11 (100) | 6/6 (100) | 7/10 (70) | 12/20 (60) |
| Propranolol | 11/27 (41) | 8/11 (72) | 1/6 (17) | 0/7 (0) | 3/12 (25) |
| Inderal LA | 1/27 (3) | 0/11 (0) | 0/6 (0) | 1/7 (14) | 0/12 (0) |
| Atenolol | 4/27 (15) | 0/11 (0) | 0/6 (0) | 0/7 (0) | 1/12 (8) |
| Nadolol | 5/27 (19) | 3/11 (27) | 4/6 (67) | 6/7 (86) | 6/12 (50) |
| Unknown | 6/27 (22) | 0/11 (0) | 1/6 (17) | 0/7 (0) | 2/12 (17) |
| Additional Medications |  |  |  |  |  |
| Mexiletine | 5/29 (17) | 3/11 (27) | 4/6 (67) | 0/8 (0) | 2/21 (10) |
| Verapamil | 1/29 (3) | 0/11 (0) | 0/6 (0) | 0/8 (0) | 1/15 (7) |
| Nifedipine | 2/29 (7) | 0/11 (0) | 0/6 (0) | 0/6 (0) | 0/6 (0) |

#### Beta-Blockers

Twenty-seven of 30 TS1 individuals receive beta-blockers (BB) for management of Long QT syndrome (LQTS). In our survey cohort, the BBs prescribed include propranolol (11/27, 41%), atenolol (4/27, 15%), and nadolol (5/27, 19%). Of the three who did not take BBs, one child could not tolerate BB medications, a second child was prescribed verapamil, and a third was prescribed mexiletine only. Five of 19 (26%) TS1 individuals take BBs combined with mexiletine for arrhythmia control. One adult TS1 individual is on BBs and nifedipine. All 11 individuals in our TS2 Gly406Arg cohort take BBs. Most are prescribed propranolol (8/11, 72%), and the remaining children take nadolol (3/11, 27%). Three TS2 Gly406Arg children also take mexiletine in addition to a BB (3/11, 27%). (Table 10: Cardiac Medications and Procedures)

Children with the Gly402Ser variant all take BBs (6/6), most commonly nadolol (4/6, 67%), and one child takes propranolol (1/6, 17%). Four of 6 (67%) are also prescribed mexiletine. For diagnosed nsLQT8 patients, BBs are also common (7/10, 70%), with 6 of 7 (86%) taking nadolol and 1 individual prescribed Inderal LA. Individuals with other variants are commonly prescribed BBs (12/20, 60%), including propranolol (3/12, 25%), atenolol (1/12, 8%), and nadolol (6/12, 50%). Two of 21 patients also take mexiletine, and 1 is prescribed verapamil.

#### Left Cardiac Stellate Denervation (LCSD)

The surgical procedure of left cardiac stellate denervation (LCSD) is sometimes undertaken to treat children with TS, despite high risk for arrhythmias under anesthesia. Typically, LCSD is performed when medication protocols are not tolerated or when uncontrolled adrenergically-driven arrhythmias are suspected. It is currently unclear if the TS or LQT8 arrhythmias are caused solely by adrenergic triggers since sleep and/or early morning arrhythmias are noted (Table 3: Early Development – Cardiac Concerns). In our surveyed cohort, 7 of 30 (23%) TS1 children, 1 of 11 (9%) TS2 Gly406Arg children, and 2 of 6 (33%) Gly402Ser children have undergone LCSD. From the literature, we are aware of one additional TS1^10^ and TS2 Gly406Arg ^13^ child who underwent this procedure (Table 10: Cardiac Medications and Procedures).

The efficacy of LCSD in survival and control of arrhythmias, according to parental survey answers, indicates mixed results. Side effects of LCSD are generally minimal, though Horner’s syndrome is a common side effect^36^. Our survey assessed LCSD efficacy in arrhythmia prevention compared to pre-surgery. TS1, TS2 Gly406Arg and Gly402Ser children were all drug compliant before LCSD surgery. Survival rates with LCSD in TS1 is currently 57% (4/7), and 43% (3/7) have died since LCSD surgery. Three of seven (43%) TS1 children were prescribed maximum medication protocols but continued to have arrhythmia concerns; all three children have continued to have ICD shocks after LCSD surgery. The remaining 57% of TS1 individuals underwent LCSD surgery for added protection unrelated to ICD shocks, however 3 children died from non-arrhythmia events. One TS1 child did not tolerate beta-blocker medications and underwent LCSD for protected care but ICD shocks continued; subsequently, a right cardiac stellate denervation (RCSD) was also performed with no further ICD shocks to date. The one TS2 Gly406Arg child who received the surgery has died as a result of respiratory arrest without arrhythmia. The two diagnosed nsLQT8 Gly402Ser children have continued to endure ICD shocks but with reduced numbers as perceived by parents.

## DISCUSSION

### Variable symptom presentation is universal among individuals with *CACNA1C* variants

A major theme emerging from the comparison of these different groups of patients is the multisystem nature of the disorder, even among patients previously diagnosed with cardiac-selective effects. While the degree and pattern of system effects differed across and within our analysis groups, it is apparent that any patient may be susceptible to a wide variety of symptoms. The Gly406Arg variant does appear most likely to cause a more severe phenotype with higher incidence of both cardiac and non-cardiac features, though some differences persist between TS1 and TS2 Gly406Arg, such as presence of syndactyly exclusive to TS1 and higher instances of bone and joint concerns in TS2 Gly406Arg. Also of note, Gly402Ser individuals differ from TS2 Gly406Arg in their lack of cardiac concerns *in utero*, leading to later recognition of the disease. However, for most categories of symptoms, Gly402Ser, nsLQT8, and other *CACNA1C* variants were often difficult to distinguish, exhibiting overlapping symptoms that varied significantly across the patient population. Thus, we propose that Timothy syndrome and *CACNA1C* related disorder present as a spectrum of symptoms, with any patient harboring a *CACNA1C* allele at potential risk for both cardiac and non-cardiac phenotypes regardless of the initial diagnosis, making the nsLQT8 diagnosis a misnomer. At the same time, the severe disease first described by Splawski et. al.^10^ represents the most severe manifestation of the disease, with many patients in this study reporting a less debilitating variation of the disease and significantly increased survival. Finally, it is important to note that several of the respondents for children with other variants did not report any cardiac features. Given that most patients have historically been identified following a cardiac event, it is probable that patients without severe cardiac effects are underrepresented in this study. Yet, it is important to note that not all patients with reported LQT presented at birth, leaving open the possibility that patients initially diagnosed with normal or borderline QT intervals may be diagnosed later in life, and may be susceptible to acquired LQT.

### Life-threatening cardiac and extra-cardiac symptoms

Our study has revealed considerable new insights on three life-threatening conditions faced by TS individuals that were not apparent when the syndrome was first described. These include: 1) cardiac arrhythmias and SCA, 2) clinically significant hypoglycemia, and 3) defects in respiratory function.

#### 1. Arrhythmia

Since the discovery of TS, LQTS and life-threatening cardiac arrhythmias have been the hallmark features. Individuals with TS are at risk of SCA due to underlying prolonged QT interval. Even those patients harboring a *CACNA1C* variant that does not present with LQT should be aware of potential cardiac risk. They also must avoid medications that further prolong QT interval to avoid risk of cardiac events^37^. All TS patients require prompt treatment; ICD implantation should be considered upon diagnosis to reduce the risk of mortality.

#### 2. Hypoglycemia

Unusual dips in blood sugar are a demonstrated concern across all groups within this study (Table 4: Hypoglycemia and Endocrine Dysfunction). Hypoglycemia has been implicated as the underlying cause of death of at least one child with TS1. Some hypoglycemic episodes have no recognizable associations and appear to be spontaneous.

Hypoglycemia occurs commonly in TS newborns, particularly those born pre-term. In our study, 5/6 (83%) of hypoglycemic TS1 newborns were born pre-term. Whether this hypoglycemia is due to TS directly, or indirectly due to the metabolic stress of delivery in combination with insufficient glycogen reserves in pre-term newborns is unknown.

#### 3. Respiratory dysfunction

Respiratory dysfunction in TS can be severe, manifesting as spontaneous respiratory arrest. One child with TS2 Gly406Arg died following documented primary pulmonary hypertension without documented arrhythmia. Respiratory associated death was also noted in 3 additional patients, one with TS1 and 2 with other variants. Recurrent pneumonias in early childhood, especially in boys with TS1, followed by asthma requiring nebulizer use, are also noted to occur. The cause of respiratory dysfunction is unknown but could be associated with compromised pulmonary development consequent to premature birth and/or to abnormal lung function that occurs with Cesarean deliveries^38–40^ which were prevalent obstetric complications in our study.

### Review of therapies and treatment options

#### 1. ICDs, AEDs, and home monitoring

Decisions regarding ICD implantation in the TS population are complicated by young age at time of diagnosis. Most patients are too small for conventional transvenous systems or a subcutaneous device, resulting in the need for modified epicardial implants which are known to have a relatively high rate of complications and need for early reintervention. In our survey, many parents expressed a desire to avoid early ICD implant and rely instead on noninvasive monitoring (e.g. Owlet sock monitor during sleep or a pulse oximeter), with a standby AED always available for the first years few of life. Implantable loop recorders^41, 42^ can be considered for arrhythmia surveillance, and for those with pacemakers, tachycardia detection algorithms can be carefully monitored, perhaps allowing early detection of non-sustained arrhythmias that may influence decisions regarding the timing of ICD implant, although neither of these modalities can provide an instantaneous alert should a sustained ventricular arrhythmia occur^43^. Close daily observation by parents and other knowledgeable caregivers (with an AED constantly nearby) remains critical for safe monitoring of TS patients without ICDs in place.

#### 2. Left Cardiac Sympathetic Denervation (LCSD)

As there are a limited number of TS children who have undergone LCSD surgery, the utility of this procedure on individuals with TS is still unclear. For TS1 children who underwent this procedure 3 of 7 have subsequently died (Table 10: Cardiac Medications and Procedures), though it is understood by ICD interrogation that 2/3 TS1 children died of non-arrhythmic causes (hypoglycemia and an extra-cardiac dysfunction). Unfortunately, it is challenging to make general recommendations for this procedure for any child with TS since the sample size is small and there have not been clinical trials with follow-up. Additionally, many parents express hesitancy about early surgical procedures, such as LCSD or ICD implantation, as anesthesia and infection can pose major risks.

#### 3. Pre-term delivery

Our study raises questions regarding the necessity of early pre-term delivery for fetuses with TS and intermittent 2:1 AVB. Traditionally, there has been a tendency to consider pre-term delivery for TS fetuses with intermittent 2:1 AVB or bradycardia under the assumption that pre-term delivery may mitigate potential risks related to bradycardia^44^. However, whether the advantages of pre-term delivery outweigh those of continued *in utero* development is an open question. Future study of developmental outcomes in infants born without early recognized cardiac manifestations may shed light on the risks related to early delivery and potentially enhance the care and management of these patients.

#### 4. Monitoring/treating hypoglycemia

Intermittent hypoglycemia has been associated with sudden death in TS children. Although this was reported in the original 2004 publication^10^, our survey has revealed that greater awareness of this phenotype is necessary. It is clear from our surveys that parents monitor their children’s glucose levels and have developed strategies to prevent hypoglycemia after an overnight fast. Despite this, more studies are needed to understand the relationship between illness/infection and low blood glucose levels.

#### 5. Awareness of anesthesia risks

Anesthesia remains a serious threat to TS individuals. Twenty-one of 30 (70%) TS1 and 8 of 11 (72%) TS2 Gly406Arg children have suffered serious arrythmias upon administration of anesthesia or during surgery for correction of syndactyly, hip alignment, or dental extractions. It remains questionable whether hypoglycemia resulting from pre-surgery fasting is linked to or exacerbates arrhythmias during anesthesia for children with TS.

#### 6. Gastro-intestinal challenges

In over 30 years of following TS children, the esophageal-gastrointestinal concerns have been under recognized, assumed to be due to a lack of GI evaluations. However, our study indicates they are among the more pressing health concerns for immediate care and survival. Esophageal concerns in infants and young children, and the development of significant GI problems as they mature were identified across all analyzed groups. This emphasizes the need for further evaluation and clarification of care specifically for the concerning effects most commonly caused by constipation in young TS children, and the deterioration of renal function associated with constipation as they age. Given the known tissue expression of *CACNA1C* in smooth muscle (www.proteinatlas.org), we hope these findings promote greater interest in GI research in the TS context.

## Conclusions

Overall, our study highlights both shared and unique symptoms among TS1, TS2 Gly406Arg, LQT8, and individuals with other *CACNA1C* variants. We find a variety of extra-cardiac symptoms, across all groups analyzed. Several of these features, such as gastrointestinal concerns and hypoglycemia, were identified in every group, suggesting that any variant in *CACNA1C* has the potential to be syndromic. We hope that our present study has brought to light the serious extracardiac phenotypes that can impact survival rate and quality of life. We believe a better understanding of the cardiac and extra cardiac concerns faced by these patients will lead to more comprehensive disease management and identify necessary avenues of TS research to improve treatments for patients.

## Data Availability

All data produced in the present work are contained in the manuscript.
Due to privacy concerns, full individualized datasets (including potentially identifiable information) are not publicly available.

## List of Abbreviations

TS: Timothy syndrome
TS1: Timothy syndrome type 1
TS2: Timothy syndrome type 2
LQT: Long QT
LQT8: Long QT type 8
nsLQT8: Non-syndromic Long QT type 8
ADD: Attention deficit disorder
ADHD: Attention-deficit/hyperactivity disorder
ICD: Implantable cardiac defibrillator
AED: automated external defibrillator
BB: Beta blocker
SUD: Sudden unexplained death
SADS: Sudden Arrythmia Death Syndromes
AVB: Atrioventricular block
PDA: Patent ductus arteriosus
PFO: Patent foramen ovale
VSD: Ventricular septal defect
HCM: Hypertrophic cardiomyopathy
ToF: Tetralogy of Fallot
CHD: Congenital heart defect
LVH: Left ventricular hypertrophy
SCA: Dudden cardiac arrest
LCSD: Left cardiac stellate denervation

## DECLARATIONS

### Ethics approval and consent to participate

Data within this study was collected after informed consent from the TS individual or parent/guardian completing the survey on their behalf. The study was approved by the Institutional Review Board of Genetic Alliance (Washington, DC, USA).

### Consent for publication

All participants provided consent consistent with the IRB approved by Genetic Alliance.

### Availability of data and material

Due to privacy concerns, the full datasets are not publicly available.

### Author’s contributions

KWT and AG conceived the study. RB and KAL collected the data and prepared the figures. KWT RB KAL IED and AG analyzed the results. KWT, RB, KAL and AG wrote the initial draft. KWT, RB, KAL, EPW, DJA, CGC, AV, GSP, IED and AG interpreted the results, edited, and revised the manuscript.

## Acknowledgements

This work was done under the direction of Dr. Andy Golden, who sadly passed away while this manuscript was in preparation. Andy was a tremendous scientist and mentor who cared deeply for improving the lives of TS patients and their families. We would also like to extend our sincere gratitude to the TS patients and families who participated in this study.

## References

1. Schulze-Bahr, E., Haverkamp, W., Borggrefe, M., Wedekind, H., Mönnig, G., Mergenthaler, J., Assmann, G., Funke, H., and Breithardt, G. (2000). Molecular genetics of arrhythmias – a new paradigm. Zeitschrift für Kardiologie 89, IV12–IV22. 10.1007/s003920070059.

2. Bokil, N.J., Baisden, J.M., Radford, D.J., and Summers, K.M. (2010). Molecular genetics of long QT syndrome. Molecular Genetics and Metabolism 101, 1–8. 10.1016/j.ymgme.2010.05.011.

3. Curran, M.E., Splawski, I., Timothy, K.W., Vincen, G.M., Green, E.D., and Keating, M.T. (1995). A molecular basis for cardiac arrhythmia: HERG mutations cause long QT syndrome. Cell 80, 795–803.

4. Wang, Q., Curran, M.E., Splawski, I., Burn, T., Millholland, J., VanRaay, T., Shen, J., Timothy, K., Vincent, G., and De Jager, T. (1996). Positional cloning of a novel potassium channel gene: KVLQT1 mutations cause cardiac arrhythmias. Nature genetics 12, 17–23.

5. Wang, Q., Shen, J., Splawski, I., Atkinson, D., Li, Z., Robinson, J.L., Moss, A.J., Towbin, J.A., and Keating, M.T. (1995). SCN5A mutations associated with an inherited cardiac arrhythmia, long QT syndrome. Cell 80, 805–811.

6. Reichenbach, H., Meister, E.M., and Theile, H. (1992). [The heart-hand syndrome. A new variant of disorders of heart conduction and syndactylia including osseous changes in hands and feet]. Kinderarztl Prax 60, 54–56.

7. Levin, S., Harrisberg, J., Govandrageloo, K., and DuPlessis, J. (1992). Idiopathic long QT syndrome in a black infant. Cardiovascular Journal of Africa 3, 144–146.

8. Marks, M.L., Trippel, D.L., and Keating, M.T. (1995). Long QT syndrome associated with syndactyly identified in females. The American Journal of Cardiology 76, 744–745. 10.1016/S0002-9149(99)80216-1.

9. Marks, M.L., Whisler, S.L., Clericuzio, C., and Keating, M. (1995). A new form of long QT syndrome associated with syndactyly. Journal of the American College of Cardiology 25, 59–64.

10. Splawski, I., Timothy, K.W., Sharpe, L.M., Decher, N., Kumar, P., Bloise, R., Napolitano, C., Schwartz, P.J., Joseph, R.M., and Condouris, K. (2004). CaV1. 2 calcium channel dysfunction causes a multisystem disorder including arrhythmia and autism. Cell 119, 19–31.

11. Clark, M.B., Wrzesinski, T., Garcia, A.B., Hall, N.A., Kleinman, J.E., Hyde, T., Weinberger, D.R., Harrison, P.J., Haerty, W., and Tunbridge, E.M. (2020). Long-read sequencing reveals the complex splicing profile of the psychiatric risk gene CACNA1C in human brain. Molecular psychiatry 25, 37–47.

12. Hu, Z., Liang, M.C., and Soong, T.W. (2017). Alternative Splicing of L-type Ca(V)1.2 Calcium Channels: Implications in Cardiovascular Diseases. Genes (Basel) 8. 10.3390/genes8120344.

13. Splawski, I., Timothy, K.W., Decher, N., Kumar, P., Sachse, F.B., Beggs, A.H., Sanguinetti, M.C., and Keating, M.T. (2005). Severe arrhythmia disorder caused by cardiac L-type calcium channel mutations. Proceedings of the National Academy of Sciences 102, 8089–8096.

14. Panagiotakos, G., Haveles, C., Arjun, A., Petrova, R., Rana, A., Portmann, T., Paşca, S.P., Palmer, T.D., and Dolmetsch, R.E. (2019). Aberrant calcium channel splicing drives defects in cortical differentiation in Timothy syndrome. Elife 8, e51037.

15. Etheridge, S.P., Bowles, N.E., Arrington, C.B., Pilcher, T., Rope, A., Wilde, A.A., Alders, M., Saarel, E.V., Tavernier, R., and Timothy, K.W. (2011). Somatic mosaicism contributes to phenotypic variation in Timothy syndrome. American journal of medical genetics Part A 155, 2578–2583.

16. Dufendach, K.A., Giudicessi, J.R., Boczek, N.J., and Ackerman, M.J. (2013). Maternal mosaicism confounds the neonatal diagnosis of type 1 Timothy syndrome. Pediatrics 131, e1991–e1995.

17. Lo-A-Njoe, S.M., Wilde, A.A., van Erven, L., and Blom, N.A. (2005). Syndactyly and long QT syndrome (CaV1. 2 missense mutation G406R) is associated with hypertrophic cardiomyopathy. Heart Rhythm 2, 1365–1368.

18. Krause, U., Gravenhorst, V., Kriebel, T., Ruschewski, W., and Paul, T. (2011). A rare association of long QT syndrome and syndactyly: Timothy syndrome (LQT 8). Clinical Research in Cardiology 100, 1123–1127.

19. Nathan, A.T., Antzelevitch, C., Montenegro, L.M., and Vetter, V.L. (2012). Case scenario: anesthesia-related cardiac arrest in a child with Timothy syndrome. Anesthesiology 117, 1117.

20. Dufendach, K.A., Timothy, K., Ackerman, M.J., Blevins, B., Pflaumer, A., Etheridge, S., Perry, J., Blom, N.A., Temple, J., and Chowdhury, D. (2018). Clinical outcomes and modes of death in Timothy syndrome: a multicenter international study of a rare disorder. JACC: Clinical Electrophysiology 4, 459–466.

21. Gettys, F.K., and Gaston, R.G. (2014). Timothy syndrome: a life-threatening syndactyly association: a case report. JBJS Case Connector 4, e48.

22. Corona-Rivera, J.R., Barrios-Prieto, E., Nieto-García, R., Bloise, R., Priori, S., Napolitano, C., Bobadilla-Morales, L., Corona-Rivera, A., Zapata-Aldana, E., Peña-Padilla, C., et al. (2015). Unusual retrospective prenatal findings in a male newborn with Timothy syndrome type 1. European Journal of Medical Genetics 58, 332–335. 10.1016/j.ejmg.2015.04.001.

23. Kawaida, M., Abe, T., Nakanishi, T., Miyahara, Y., Yamagishi, H., Sakamoto, M., and Yamada, T. (2016). A case of Timothy syndrome with adrenal medullary dystrophy. Pathology international 66, 587–592.

24. Sepp, R., Hategan, L., Bácsi, A., Cseklye, J., Környei, L., Borbás, J., Széll, M., Forster, T., Nagy, I., and Hegedűs, Z. (2017). Timothy syndrome 1 genotype without syndactyly and major extracardiac manifestations. American Journal of Medical Genetics Part A 173, 784–789.

25. Walsh, M.A., Turner, C., Timothy, K.W., Seller, N., Hares, D.L., James, A.F., Hancox, J.C., Uzun, O., Boyce, D., and Stuart, A.G. (2018). A multicentre study of patients with Timothy syndrome. EP Europace 20, 377–385.

26. Gao, Y., Xue, X., Hu, D., Liu, W., Yuan, Y., Sun, H., Li, L., Timothy, K.W., Zhang, L., Li, C., and Yan, G.-X. (2013). Inhibition of Late Sodium Current by Mexiletine. Circulation: Arrhythmia and Electrophysiology 6, 614–622. doi:10.1161/CIRCEP.113.000092.

27. Rios, R., Wakai, R.T., and Strasburger, J.F. (2015). A rare congenital long QT syndrome diagnosed prenatally with fetal magnetocardiography. Neonatology Today 10, 1–6.

28. An, H.S., Choi, E.Y., Kwon, B.S., Kim, G.B., Bae, E.J., Noh, C.I., Choi, J.Y., and Park, S.S. (2013). Sudden cardiac arrest during anesthesia in a 30-month-old boy with syndactyly: a case of genetically proven Timothy syndrome. Journal of Korean medical science 28, 788–791.

29. Fukuyama, M., Wang, Q., Kato, K., Ohno, S., Ding, W.-G., Toyoda, F., Itoh, H., Kimura, H., Makiyama, T., Ito, M., et al. (2014). Long QT syndrome type 8: novel CACNA1C mutations causing QT prolongation and variant phenotypes. EP Europace 16, 1828–1837. 10.1093/europace/euu063.

30. Diep, V., and Seaver, L.H. (2015). Long QT syndrome with craniofacial, digital, and neurologic features: Is it useful to distinguish between Timothy syndrome types 1 and 2? American Journal of Medical Genetics Part A 167, 2780–2785.

31. Philipp, L.R., and Rodriguez III, F.H. (2016). Cardiac arrest refractory to standard intervention in atypical Timothy syndrome (LQT8 type 2). In 2. (Taylor & Francis), pp. 160–162.

32. Borbás, J., Vámos, M., Hategan, L., Hanák, L., Farkas, N., Szakács, Z., Csupor, D., Tél, B., Kupó, P., Csányi, B., et al. (2022). Geno- and phenotypic characteristics and clinical outcomes of CACNA1C gene mutation associated Timothy syndrome, “cardiac only” Timothy syndrome and isolated long QT syndrome 8: A systematic review. Frontiers in Cardiovascular Medicine 9. 10.3389/fcvm.2022.1021009.

33. Ty, J., Timothy K, Golden A, See V, and D., H. (in press). Syndactyly Patterns in Timothy Syndrome Type 1. Journal of Hand Surgery.

34. Unique (2006). 4q Deletions From 4q31and Beyond. In E.-M.H. Strehle, Maj, ed. Rare Chromosome & Gene Disorder Guides. Unique.

35. Agarwal, D., Karmazyn, B., Corea, D., and Kaefer, M. (2013). Pediatric penile pain secondary to idiopathic arterial thrombosis. Journal of Pediatric Surgery Case Reports 1, 319–320. 10.1016/j.epsc.2013.08.012.

36. Cho, Y. (2016). Left cardiac sympathetic denervation: an important treatment option for patients with hereditary ventricular arrhythmias. Journal of Arrhythmia 32, 340–343.

37. Kallergis, E.M., Goudis, C.A., Simantirakis, E.N., Kochiadakis, G.E., and Vardas, P.E. (2012). Mechanisms, risk factors, and management of acquired long QT syndrome: a comprehensive review. The Scientific World Journal 2012.

38. Colin, A.A., McEvoy, C., and Castile, R.G. (2010). Respiratory morbidity and lung function in preterm infants of 32 to 36 weeks’ gestational age. Pediatrics 126, 115–128.

39. Reuter, S., Moser, C., and Baack, M. (2014). Respiratory distress in the newborn. Pediatrics in review 35, 417–429.

40. Steinhorn, R.H. (2010). Neonatal pulmonary hypertension. Pediatr Crit Care Med 11, S79–84. 10.1097/PCC.0b013e3181c76cdc.

41. Czosek, R.J., Zang, H., Baskar, S., Anderson, J.B., Knilans, T.K., Ollberding, N.J., and Spar, D.S. (2021). Outcomes of Implantable Loop Monitoring in Patients <21 Years of Age. Am J Cardiol 158, 53–58. 10.1016/j.amjcard.2021.07.034.

42. Davidson, S., Wong, A., and Cortez, D. (2023). Loop recorder implantation in patients as young as 3-months of age; the benefit of the sub-scapular approach. Pacing Clin Electrophysiol 46, 1073–1076. 10.1111/pace.14802.

43. Bezzerides, V.J., Walsh, A., Martuscello, M., Escudero, C.A., Gauvreau, K., Lam, G., Abrams, D.J., Triedman, J.K., Alexander, M.E., Bevilacqua, L., and Mah, D.Y. (2019). The Real-World Utility of the LINQ Implantable Loop Recorder in Pediatric and Adult Congenital Heart Patients. JACC Clin Electrophysiol 5, 245–251. 10.1016/j.jacep.2018.09.016.

44. Matthews, A., Timothy, K., Golden, A., and Gonzalez Corcia, M.C. (2024). International Cohort of Neonatal Timothy Syndrome. Neonatology, 1-8. 10.1159/000535221.

